# Body Mass Index (BMI) and BMI Variability are Risk Factors for Heart Failure with Preserved and Reduced Ejection Fraction in a Longitudinal Cohort Study Using Real-World Electronic Health Records

**DOI:** 10.1101/2023.10.16.23297111

**Authors:** Zeshui Yu, Yuqing Chen, Manling Zhang, Ning Feng, Tim P Ryan, Nanette Cathrin Schloot, Yu Chen, Flora Sam, Lirong Wang

**Author notes:** Corresponding Authors: Yu Chen, PhD, Lilly Research Laboratories, Eli Lilly and Company, Lilly Corporate Center, Indianapolis, IN, 46225; Flora Sam, MD, Lilly Research Laboratories, Eli Lilly and Company Lilly Corporate Center, Indianapolis, IN, 46225; Lirong Wang, PhD, Department of Pharmaceutical Sciences, Computational Chemical Genomic Screening Center, University of Pittsburgh School of Pharmacy, Pittsburgh, PA 15213.

## Abstract

**Objective:** The aim of this study is to evaluate the differential impact of BMI and long-term intra-individual BMI variability on the risk of developing heart failure with preserved ejection fraction (HFpEF) and heart failure with reduced ejection fraction (HFrEF)in overweight or obese patients.

**Method:** The primary outcome was the time to incident HFrEF or HFpEF determined by International Classification of Disease codes (ICD-9 and ICD-10). BMI variability was assessed based on five-year BMI measurements using four metrics: the intra-individual standard deviation (SD), the coefficient of variation (CV), the variability independent of the mean (VIM), and the average successive variability (ASV). The subclassification of HF was based on the LVEF recorded within 90 days of the initial diagnosis. The hazard ratios (HRs) were estimated by multivariable-adjusted Cox proportional hazards regression models.

**Results:** Among the 51,444 eligible patients, 1,871 developed HFpEF, and 1,018 developed HFrEF over a follow-up period of the mean of 4.62 years. Per each 1-SD increment, the HRs of SD, CV, VIM and ASV of BMI were 1.10 (95%CI, 1.04-1.16), 1.04 (95%CI, 1.02-1.06), 1.16 (95%CI, 1.06-1.27) and 1.13 (95%CI, 1.04-1.22) for HFpEF, and 1.09 (95%CI, 1.00-1.18), 1.03 (95%CI, 1.00-1.06), 1.15 (95% CI, 1.02-1.31), and 1.08 (95%CI, 0.96-1.21), for HFrEF, respectively. Five statistical models were performed adjusting for different sets of covariates. Moreover, baseline BMI from patients in obesity II and III all exhibited progressively higher HRs for HFpEF with HRs of 1.45 (95%CI, 1.15-1.83), and 2.52 (95%CI, 1.99-3.19), while only patients in obesity class III demonstrated an increased risk for HFrEF with HR of 1.50 (95%CI, 1.11-2.04).

**Conclusions and Relevance:** In this large cohort of overweight and obese patients, increasing BMI variability was associated with a higher risk of developing HFpEF and HFrEF after adjusting for relevant risk factors.

**Study Importance:** **What is already known:**

- The variability of body mass index (BMI) is a risk factor for negative cardiovascular outcomes in various cohorts.
- A previous study showed that variability in adiposity indices is related to an increased risk of overall heart failure (HF) in patients with type two diabetes.

**What are the new findings in your manuscript:**

- ur analysis examined the differential association between intra-individual BMI variability derived and the increased incidence of two subtypes of (HF), heart failure with preserved ejection fraction (HFpEF) and heart failure with reduced ejection fraction (HFrEF), using real-world clinical patient data.
- associations were statistically significant across three different metrics, including standard deviation, coefficient of variation, and the variability independent of the mean.
- might your results change the direction of research or the focus of clinical practice?
- is important for clinicians to minimize body weight fluctuation in patients with overweight and obesity to meet weight loss goals due to its potential to reduce the risk of HF, especially given the increasing global prevalence of HFpEF and limited therapeutical options for it.
- study also provides evidence of the feasibility and reliability of using electronic medical data collected from various clinical settings to define indicators for clinical decision-making

## Introduction

Heart failure (HF) is a complex, progressive, and debilitating cardiovascular condition that occurs when the heart cannot efficiently pump or deliver an insufficient amount of oxygen-rich blood to meet the metabolic demands of the body. Despite some therapeutic progress, the prevalence and incidence of HF in the United States continue to rise, affecting approximately 6.2 million adults and leading to an estimate of 960,000 new cases annually.^1,2^ Heart failure is commonly classified into two major subtypes based on the measurement of left ventricular ejection fraction (LVEF), heart failure with preserved ejection fraction (HFpEF), and heart failure with reduced ejection fraction (HFrEF). Patients diagnosed with HFpEF account for at least 50% of the overall heart failure population, with the proportion of HFpEF relative to HFrEF increasing at a concerning rate of 1% per year. ^3–5^

The relationship between heart failure and obesity is complex and multifactorial and not entirely clear. The American College of Cardiology/American Heart Association guidelines acknowledges obesity as a risk factor for the development of overall HF. The latter is a composite of HFpEF and HFrEF.^6,7^ Several studies have also found that obesity portends a greater risk of developing incident HFpEF compared to HFrEF.^8,9^ However, it is important to note that the current HF guidelines do not provide definitive recommendations for weight reduction in individuals with heart failure. This is possibly because some survival benefits are seen with obesity in patients with established HF.^10,11^ Additionally, although there are multiple guideline-directed medical therapies (GDMT) that have demonstrated improved symptoms and survival in patients with HFrEF, the choice of therapy is limited in patients with HFpEF, reflecting an incomplete understanding of HFpEF.^12^

Given the current obesity epidemic, recent attention has focused on variability in body mass index (BMI) as a potential predictor of cardiovascular diseases, partly due to the common phenomenon of weight cycling, defined as intentional weight loss followed by unintentional weight regain.^13^ It is increasingly evident that fluctuation in body weight leads to adverse cardiovascular outcomes in various cohorts.^9,14–18^ While there were a few studies analyzing the association between BMI variability with incident HF, sparse data exist on a cohort of patients with overweight and obesity. ^19,20^ Additionally, this does not refer to the weight cycling that is seen in symptomatic HF where the latter refers to weight fluctuations due to fluid retention and loss with diuretics.^21^ Limited studies have investigated the possible heterogeneity of the association between variability in BMI with the incidence of HFpEF and HFrEF primarily due to the lack of imaging data, specifically echocardiography (ECHO).

Thus, the clinical utility of BMI and its variability remains to be evaluated with respect to the subtypes of HF, namely HFpEF and HFrEF. Additionally, it is unclear if previous findings can be replicated using real-world clinical data. The widespread adoption of electronic health records has offered a greater opportunity and our present hypothesis is to examine the differential risks associated with obesity severity and BMI variability for incident HFpEF and HFrEF. We conducted a retrospective cohort study with two main objectives: 1) to validate the varying risk levels of developing HFpEF and HFrEF associated with different baseline obesity levels, and 2) to investigate the association between BMI variability and incident HFpEF and HFrEF in a large cohort of overweight or obese patients diagnosed initially in the hospital settings.

## Method

### Study design

The University of Pittsburgh Medical Center (UPMC) is one of the largest healthcare systems in the United States and it provides real-world electronic health record (EHR) data for translational research.

We performed a retrospective cohort analysis by randomly selecting 100,000 patients in the UPMC healthcare system who had an initial BMI equal to or greater than 27 kg/m^2^ on presentation and were followed for at least 5 years between January 1^st^,2004, and October 31^st^, 2021. Of these individuals, 25,000 patients were each categorized into the following 4 groups based on their initial BMI values: Overweight group (27 kg/m^2^<=BMI<30 kg/m^2^), class I obesity group (30 kg/m^2^<=BMI<35 kg/m^2^), class II obesity (35 kg/m^2^<=BMI<40 kg/m^2^) and class III obesity group (BMI>=40 kg/m^2^), respectively. The BMI variability measurement period encompassed a five-year span following the date of the initial BMI value, during which BMI records documented in EHR were used to create an independent variable representing the variability in BMI for each patient. The baseline was defined as the end of the BMI variability measurement period, from which the incident HF was then evaluated. The study design is shown in Figure 1.

**Figure 1.**
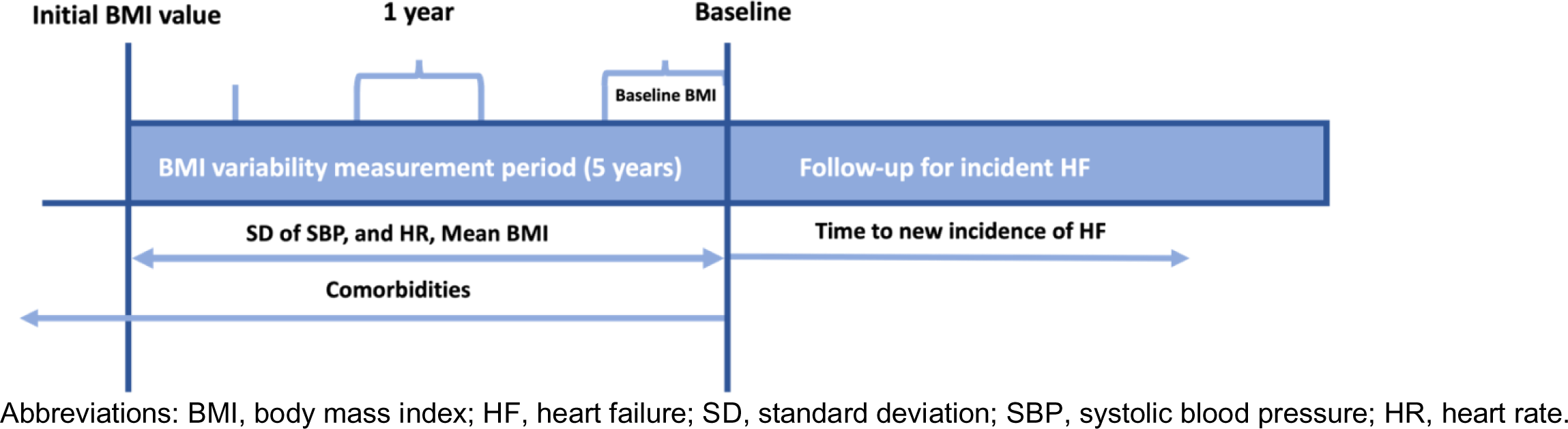
Study Design. Abbreviations: BMI, body mass index; HF, heart failure; SD, standard deviation; SBP, systolic blood pressure; HR, heart rate.

### Inclusion/Exclusion criteria

We implemented several exclusion criteria in our analysis to ensure the validity and reliability of our findings. Specifically, patients with a history of bariatric surgery (n=2,643) were excluded using Current Procedural Terminology (CPT) code due to the significant and rapid weight loss associated with this procedure. To ensure that the patients were under active and continuous BMI measurement, they were required to have at least one record of BMI each year during the BMI variability measurement period. Patients were excluded if there were missing BMI values in any year (n=18,559). Furthermore, patients with a history of any malignant tumor were excluded at any point up until the baseline (n=11,664), as well as those with missing values for systolic, diastolic blood pressure, and heart rate (n=2,723), and those with follow-up periods of <5 years due to data errors (n=2,611). Patients who developed HF prior to the end of the 5-year assessment period (i.e., before the baseline) were also excluded (n=8,900) to ensure that the assessment of BMI variability preceded the outcomes. Patients without ECHO measurements within 90 days from the date of their first HF diagnosis were excluded (n=1,456). After implementing these exclusions, the final cohort for analysis comprised 51,444 patients. Figure 2 is the detailed flow diagram describing the exclusion criteria.

**Figure 2.**
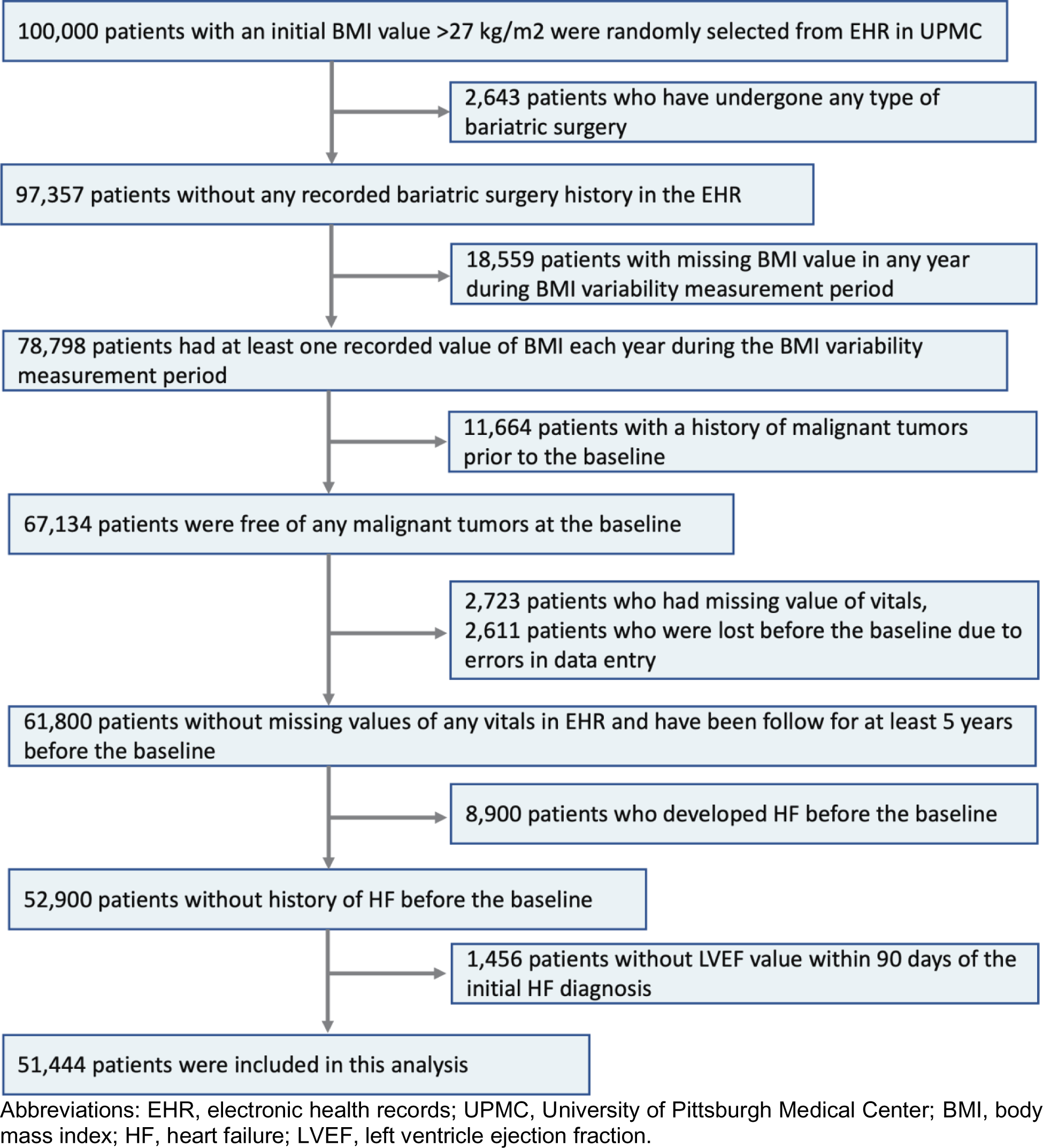
Flow Diagram for Included Patients. Abbreviations: EHR, electronic health records; UPMC, University of Pittsburgh Medical Center; BMI, body mass index; HF, heart failure; LVEF, left ventricle ejection fraction.

### Measurement of variability in BMI

Two data cleaning steps were applied to the vital records to guarantee the quality and accuracy of the BMI records used in our analysis. First, vital records with extreme BMI values that fell below 10 kg/m^2^ or above 65 kg/m^2^ were removed to ensure a reasonable and valid range of BMI. Second, since BMI values were rounded to two decimal points in the EHR system, the records with the exact same value as the previous data were excluded to account for potential duplicate values resulting from data entry errors.

BMI variability was assessed during the five-year BMI variability measurement period using four different metrics. These included the intra-individual standard deviation (SD), the coefficient of variation (CV) calculated as 100 times the SD divided by the mean, the variability independent of the mean (VIM) calculated as 100 times the SD divided by the mean raised to the power of β, which is derived from the natural logarithm of SD on the natural logarithm of the mean value of the obesity measure, and the average successive variability (ASV) calculated as the mean of the absolute difference between successive values.^16^

### The incident heart failure diagnosis

The endpoint assessed in this study is the incidence of new-onset HF, which is defined as the first-time diagnosis of clinical HF ascertained by the presence of ICD-9 and ICD-10 codes during the follow-up. The follow-up period was defined as the time interval from the baseline to the earliest occurrence of HF, death, the last encounter date, or the end of follow-up (October 31st, 2021), whichever came first. Furthermore, ECHO reports of all patients were collected and LVEF value from the most recent ECHO test conducted within 90 days of the date of the first HF diagnosis was used to determine the LVEF in patients with incident HF. HFpEF is defined as LVEF > 50%, while HFrEF is defined as LVEF ≤ 50%.^22^

### Covariates

In this study, several covariates were included to account for potential confounding factors. These covariates included age, gender, race, smoking status, the number of BMI records during the measurement period, baseline BMI categories, and comorbidities including atherosclerotic cardiovascular disease, type 2 diabetes mellitus, hypertension, hyperlipidemia, chronic kidney disease, nonalcoholic fatty liver disease/nonalcoholic steatohepatitis, and obstructive sleep apnea. Mean BMI and standard deviation of heart rate and systolic blood pressure during the BMI variability measurement period were determined. The selection of covariates was based on the previously reported potential risk factors for HF. In addition, the delta BMI changes over the BMI variability measurement period were calculated by measuring the difference between the initial BMI and the baseline BMI.

Several EHR data elements were reviewed to compile the variables for this study. Each patient’s demographic information was determined based on the record closest to the date when the initial BMI was collected. The baseline BMI value was defined as the most proximal BMI value to the baseline, which was collected within 1 year prior to the baseline according to our study design. Comorbidities were identified based on ICD-9 and ICD-10 codes documented clinical diagnosis prior to the baseline. The mean and standard deviation of vitals, such as systolic blood pressure, diastolic blood pressure, and heart rate, were calculated using the values assessed during the BMI variability measurement period. Smoking status was collected during this period using social history files.

### Statistical analysis

Patient characteristics were summarized as mean ± standard deviation for continuous variables and count (percent) for categorical variables. The within-patient BMI variability was calculated using all four metrics. We compared the distribution of the baseline demographics and characteristics by baseline BMI classes and by quartiles of CV of BMI, separately, using Chi-square test (categorical variables) and ANOVA (continuous variables). Multivariable Cox regression analysis was performed to investigate the association between 5-year BMI variability and the risk of incident heart failure during the follow-up period. The incidence rates of HF were computed as the ratio of the total number of patients diagnosed with first-time HF to the total person-years of follow-up. The baseline BMI values were classified into five groups based on the following range <27,27-30,30-35,35-40,>40 kg/m^2^. Each BMI variability metric was modeled as both continuous and categorical variables (quartiles of each BMI variability metric calculated from the entire study sample). The lowest quartile served as the reference category.

Initially, to examine if five-year BMI variability is differentially associated with the risk of developing each type of HF, Cox proportional hazard models with cause-specific competing risks analysis were applied to investigate the association of BMI variability with relative hazard ratios (HRs) on the incidence of the one type of HF by treating the other type as a competing risk. By accounting for the other type of HF as a censored event in the model, the adjusted HRs of one type of HF in the presence of the other type were reported. For each BMI variability metric, five models were performed, in model 1, age, gender, race, smoking status, and the number of BMI records during the variability measurement phase were included. Then, baseline BMI categories were further adjusted in model 2 and comorbidities in model 3. Furthermore, the SD of systolic blood pressure and heart rate were added in model 4. Finally, when the variability was assessed using SD, CV, and ASV, we further adjusted for mean BMI in addition to model 4 (model 5).

Secondly, similar Cox proportional hazard regressions for risk analysis were applied to examine the impact of baseline BMI categories on the incidence of HFpEF and HFrEF. For each type of HF, two separate models were implemented. In model 1, age, gender, race, smoking status, and the number of BMI records during the variability measurement phase were adjusted. In model 2, comorbidities were further added in model 1.

A sensitivity analysis was conducted to address potential biases associated with prolonged time intervals between the assessment of BMI variability and the incidence of HF. This analysis involved restricting the maximum follow-up duration to 7.5 years, approximating the 90th percentile of the total follow-up period. This analysis aims to mitigate potential confounding effects caused by extended time gaps.

Statistical significance was defined as a two-sided p-value less than 0.05. All statistical analyses were performed using Python 3.9 and SAS

## Results

### Characteristics of Study Patients

During the mean follow-up period of 4.62 years for the entire study patients, a total of 2,889 patients developed a new onset of heart failure, with 1,871 cases classified as HFpEF and 1,018 cases classified as HFrEF. Table 1 displays the baseline demographics and clinical characteristics of the study sample by the quartiles of CV of BMI. In comparison with patients with the lower quartile of BMI variability, patients with a higher quartile were younger and more likely to be female and black. The proportion of T2DM, CKD, NAFLD/NASH, and OSA increased across increasing quartiles of BMI variability. Interestingly, it is worth noting that there was no significant difference in the proportion of ASCVD across the quartiles of BMI variability. Furthermore, the proportion of HTN and HLP demonstrated a decreasing trend as quartiles increased. Patients in the fourth quartile also had higher means of heart rate and BMI, higher SDs of heart rate, SBP, and DBP, and lower mean SBP and DBP compared to those in the first quartile. Patients in the third quartile of the CV of BMI experienced the highest magnitude of delta weight gain over the five-year BMI measurement phase, as compared to patients in the other quartiles. Differences in baseline variables of the study sample based on baseline BMI category and HF subtype were provided in eTables 1 and 2 in the Supplement.

**Table1.** Baseline Demographics and Characteristics by Quartiles of Coefficient of Variation of BMI.

| Characteristics |  | Study Patients | Q1 | Q2 | Q3 | Q4 | P-Value |
| --- | --- | --- | --- | --- | --- | --- | --- |
| n |  | 51444 | 12861 | 12861 | 12861 | 12861 |  |
| Age, mean (SD) |  | 52.83 (14.66) | 56.61 (12.51) | 54.21 (13.68) | 52.15 (14.93) | 48.34 (16.02) | <0.001 |
| Gender, n (%) | FEMALE | 31408 (61.05) | 6592 (51.26) | 7412 (57.63) | 8196 (63.73) | 9208 (71.60) | <0.001 |
|  | MALE | 20036 (38.95) | 6269 (48.74) | 5449 (42.37) | 4665 (36.27) | 3653 (28.40) |  |
| Race, n (%) | WHITE | 44521 (86.54) | 11497 (89.39) | 11247 (87.45) | 11103 (86.33) | 10674 (83.00) |  |
|  | BLACK | 6257 (12.16) | 1214 (9.44) | 1441 (11.20) | 1605 (12.48) | 1997 (15.53) | <0.001 |
|  | OTHER | 666 (1.29) | 150 (1.17) | 173 (1.35) | 153 (1.19) | 190 (1.48) |  |
| Smoking status, mean (SD) |  | 6610 (12.85) | 1161 (9.03) | 1486 (11.55) | 1804 (14.03) | 2159 (16.79) | <0.001 |
| Num of BMI records, mean (SD) |  | 19.69 (10.86) | 15.84 (7.63) | 18.67 (9.55) | 20.60 (11.07) | 23.65 (12.92) | <0.001 |
| Baseline BMI Categories, n (%) | <27 | 2205 (4.29) | 332 (2.58) | 447 (3.48) | 607 (4.72) | 819 (6.37) | <0.001 |
|  | Overweight | 10971 (21.33) | 3763 (29.26) | 3021 (23.49) | 2383 (18.53) | 1804 (14.03) |  |
|  | Obesity I | 14454 (28.10) | 3673 (28.56) | 3752 (29.17) | 3683 (28.64) | 3346 (26.02) |  |
|  | Obesity II | 12250 (23.81) | 3047 (23.69) | 3114 (24.21) | 3174 (24.68) | 2915 (22.67) |  |
|  | Obesity III | 11564 (22.48) | 2046 (15.91) | 2527 (19.65) | 3014 (23.44) | 3977 (30.92) |  |
| ASCVD, n (%) |  | 10540 (20.49) | 2723 (21.17) | 2636 (20.50) | 2592 (20.15) | 2589 (20.13) | 0.133 |
| T2DM, n (%) |  | 15850 (30.81) | 3724 (28.96) | 3943 (30.66) | 4103 (31.90) | 4080 (31.72) | <0.001 |
| HTN, n (%) |  | 36908 (71.74) | 9606 (74.69) | 9457 (73.53) | 9114 (70.87) | 8731 (67.89) | <0.001 |
| HLP, n (%) |  | 36862 (71.65) | 10066 (78.27) | 9653 (75.06) | 9079 (70.59) | 8064 (62.70) | <0.001 |
| CKD, n (%) |  | 4373 (8.50) | 940 (7.31) | 1044 (8.12) | 1146 (8.91) | 1243 (9.66) | <0.001 |
| NAFLD/NASH, n (%) |  | 4289 (8.34) | 875 (6.80) | 1060 (8.24) | 1129 (8.78) | 1225 (9.52) | <0.001 |
| OSA, n (%) |  | 11248 (21.86) | 2404 (18.69) | 2810 (21.85) | 2960 (23.02) | 3074 (23.90) | <0.001 |
| Mean HR, mean (SD) |  | 77.09 (8.57) | 75.35 (8.34) | 76.62 (8.41) | 77.42 (8.41) | 78.99 (8.71) | <0.001 |
| Mean SBP, mean (SD) |  | 129.21 (9.40) | 130.10 (9.30) | 129.64 (9.19) | 129.10 (9.42) | 128.01 (9.56) | <0.001 |
| Mean DBP, mean (SD) |  | 78.64 (5.75) | 78.98 (5.74) | 78.91 (5.67) | 78.62 (5.68) | 78.06 (5.86) | <0.001 |
| SD of heart rate, mean (SD) |  | 9.16 (3.43) | 8.48 (3.29) | 8.94 (3.31) | 9.26 (3.36) | 9.96 (3.59) | <0.001 |
| SD of SBP, mean (SD) |  | 11.95 (3.86) | 11.55 (3.81) | 11.86 (3.75) | 12.08 (3.82) | 12.33 (4.03) | <0.001 |
| SD of DBP, mean (SD) |  | 7.74 (2.14) | 7.36 (2.16) | 7.61 (2.10) | 7.82 (2.09) | 8.17 (2.15) | <0.001 |
| Mean BMI, mean (SD) |  | 35.30 (6.45) | 34.10 (5.88) | 34.81 (6.16) | 35.54 (6.43) | 36.76 (6.98) | <0.001 |
| Delta BMI, mean (SD) |  | 0.23 (2.75) | 0.14 (1.20) | 0.25 (1.83) | 0.33 (2.52) | 0.20 (4.37) | <0.001 |

Abbreviations: BMI, body mass index; HF, heart failure; HFpEF, heart failure with preserved ejection fraction; HFrEF, heart failure with rejected ejection fraction; SD, standard deviation; CV, coefficient of variation; VIM, variability independent of mean; ASV, average successive variability; ASCVD, atherosclerotic cardiovascular disease; T2DM, type 2 diabetes mellitus; HTN, hypertension; HLP, hyperlipidemia; CKD, chronic kidney disease; NAFLD/NASH, nonalcoholic fatty liver disease/nonalcoholic steatohepatitis; OSA, obstructive sleep apnea; SBP, systolic blood pressure; DBP, diastolic blood pressure;
Continuous variables are presented as mean values with standard deviations, while categorical variables are expressed as a proportion of patients.

### Variability of BMI and incident HF

Table 2 presents the adjusted HRs for the for incident HFpEF and HFrEF by different metrics of BMI variability. When evaluated as a continuous variable, increasing BMI variability measured by SD, CV, and VIM was positively associated with increased risk of developing HFpEF and HFrEF across all five models, after adjusting for all relevant covariates. The fully adjusted HRs for each 1-SD increment in SD, CV, and VIM of BMI were 1.10 (95%CI, 1.04-1.16), 1.04 (95%CI, 1.02-1.06), and 1.16 (95%CI, 1.06-1.27), respectively, for HFpEF, and 1.09 (95%CI, 1.00-1.18), 1.03 (95%CI, 1.00-1.06), 1.15 (95% CI, 1.02-1.31), respectively, for HFrEF (model 5). For ASV of BMI, the significant association was only observed in HFpEF. The HR for each 1-SD increment in ASV of BMI is 1.13 (95%CI, 1.04-1.22), for HFpEF (model 5).

**Table 2:**
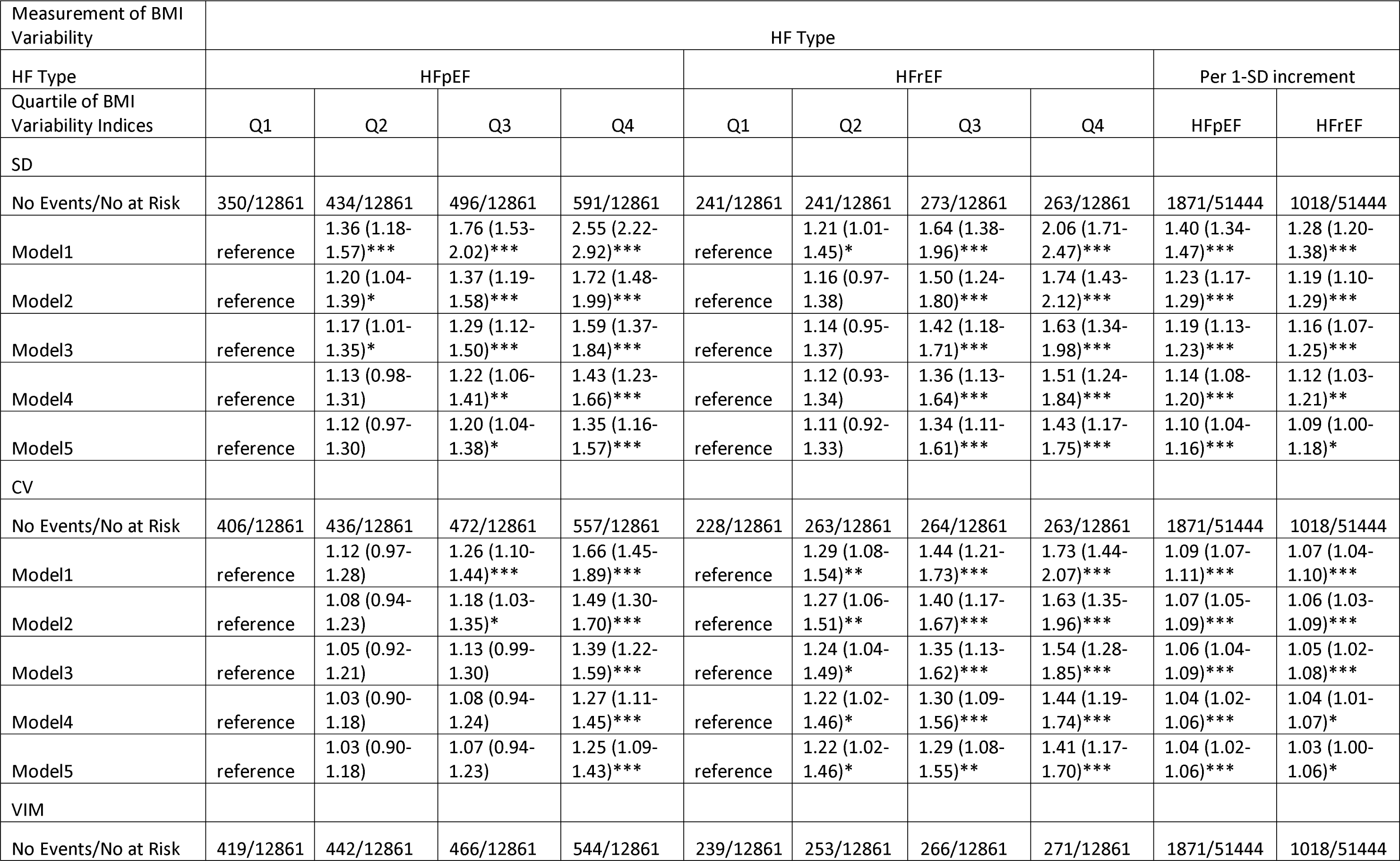
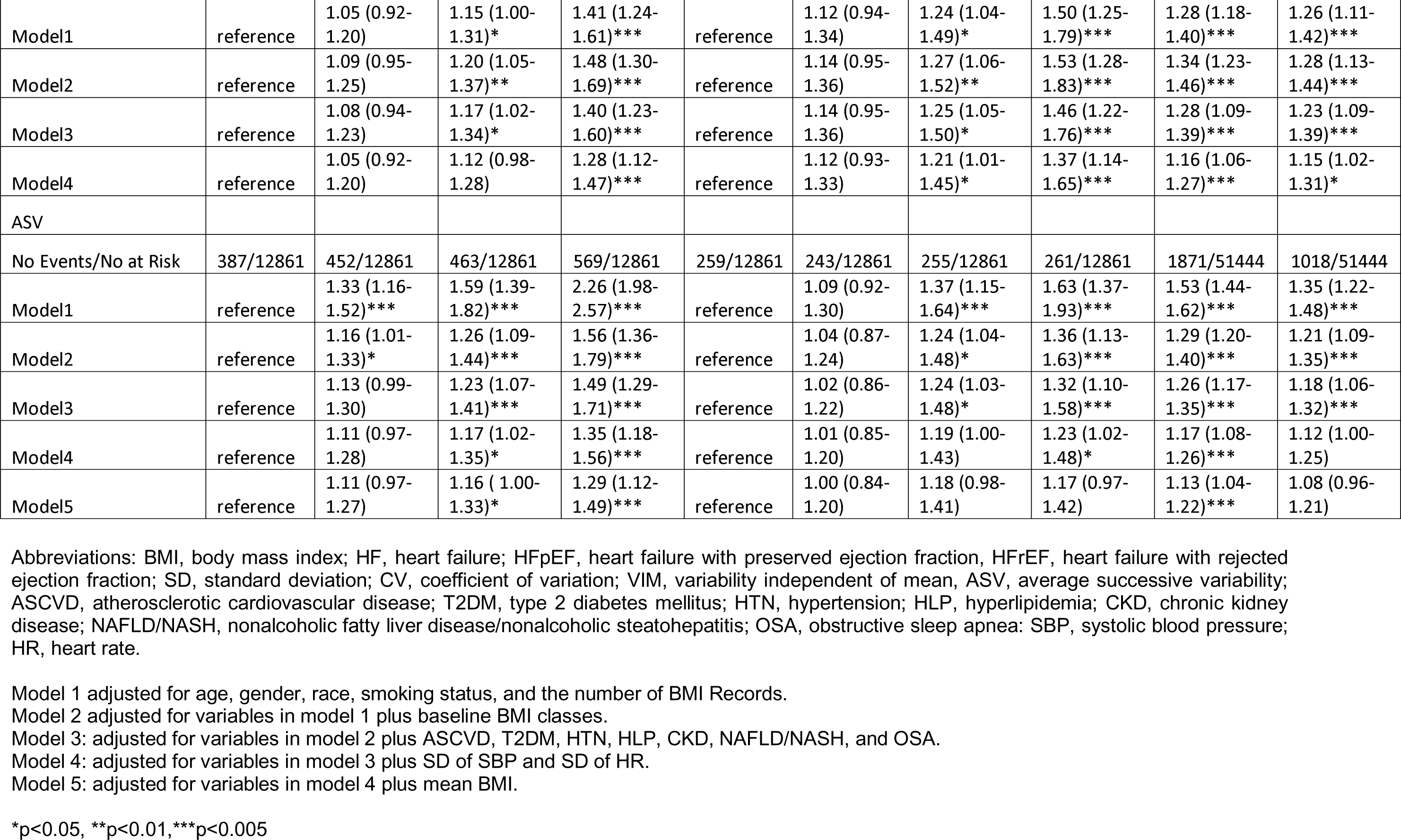
Hazard Ratios for Incident Heart Failure by BMI Variability in the Study Cohort.

When BMI variability was modeled in quartile, patients in the third and fourth quartiles of SD of BMI demonstrated increased risks for HFpEF (adjusted HR of 1.20 (95%CI, 1.04-1.38) in the third quartile and 1.35 (95%CI, 1.16-1.57) in the fourth quartile) and HFrEF (adjusted HR of 1.34 (95%CI, 1.11-1.61) in the third quartile and 1.43 (95%CI, 1.17-1.75) in the fourth quartile) compared to those in the first quartile. Additionally, the adjusted HR for the fourth quartile of CV of BMI was 1.25 (95%CI, 1.09-1.43) for HFpEF as compared to the first quartile. Patients with a CV of BMI in the second quartile and above had an increased risk for HFrEF (HRs of 1.22 (95%CI, 1.02-1.46), 1.29 (95%CI, 1.08-1.55), and 1.41 (95%CI, 1.17-1.70) for the second, third, and fourth quartiles, respectively). For VIM of BMI, the adjusted HRs for patients in the top quartile compared to the bottom quartile were 1.28 (95%CI, 1.12-1.47) for HFpEF, while for HFrEF, the HRs of elevated VIM of BMI in the third quartile and the fourth quartile were 1.21 (95%CI, 1.01-1.45), and 1.37 (95%CI, 1.14-1.65), respectively in comparison with those in the first quartile. For ASV of BMI, the HRs associated with incident HFpEF is significant in the third (HR: 1.16 (95%CI, 1.00-1.33)) and fourth quartile (HR: 1.29 (95%CI, 1.12-1.49)).

The model details relating to all selected covariates and incident HF by BMI variability metrics were shown in eTables 3, 4, 5, and 6 in the Supplement.

### Baseline BMI Class and incident HF

The adjusted HRs of the baseline BMI categories for incident HFpEF and HFrFE were presented in Table 3 for two models. Compared to patients with baseline BMI<27, patients with baseline BMI falling into the obesity class I, II and III demonstrated an elevated risk for HFpEF (Model 1; HR 1.26 (95%CI, 1.10-1.58), 1.73 (95%CI, 1.37-2.18), and 3.24 (95%CI, 2.57-4.08) for obesity class I, II, III, respectively). Despite adjusting for various chronic comorbidities, patients in obesity class II and III exhibited an increased risk for HFpEF compared with those with baseline BMI<27 (Model 2; HR 1.45 (95%CI, 1.15-1.83) and 2.52 (95%CI, 1.99-3.19) for obesity class II and III). However, it was found that only patients with severe obesity (BMI>40) were associated with a higher risk of HFrEF in both models compared to those with BMI< 27 (HR 1.89 (95%CI, 1.40-2.55) for model 1 and 1.50 (95%CI, 1.11-2.04) for model 2). Regardless, in the patients with the greatest level of obesity, ie. obesity class III, the risk is significantly worse for HFpEF compared with HFrEF.

**Table 3:** Baseline BMI class and the incidents of HFpEF and HFrEF.

| HF Type | HFpEF |  |  |  |  | HFrEF |  |  |  |  |
| --- | --- | --- | --- | --- | --- | --- | --- | --- | --- | --- |
| Baseline BMI Categories | <27 | Overweight | Obesity Class I | Obesity Class II | Obesity Class III | <27 | Overweight | Obesity Class I | Obesity Class II | Obesity Class III |
| No Events/No at Risk | 87/2205 | 285/10971 | 484/14454 | 458/12250 | 557/10793 | 57/2205 | 237/10971 | 273/14454 | 237/12250 | 214/10793 |
| Model1 | Reference | 0.82 (0.64-1.04) | 1.26 (1.00-1.58)* | 1.73 (1.37-2.18)*** | 3.24 (2.57-4.08)*** | Reference | 0.92 (0.70-1.24) | 1.10 (0.75-1.33) | 1.31 (0.98-1.76) | 1.89 (1.40-2.55)*** |
| Model2 | Reference | 0.81 (0.64-1.04) | 1.16 (0.92-1.46) | 1.45 (1.15-1.83)** | 2.52 (1.99-3.19)*** | Reference | 0.93 (0.69-1.24) | 0.91 (0.68-1.21) | 1.09 (0.81-1.47) | 1.50 (1.11-2.04)** |

Abbreviations: BMI, body mass index; HF, heart failure; HFpEF, heart failure with preserved ejection fraction; HFrEF, heart failure with rejected ejection fraction.
Model 1 adjusted for age, gender, race, smoking status, and the number of BMI Records.
Model 2 adjusted for variables in model 1 plus ASCVD, T2DM, HTN, HLP, CKD, NAFLD/NASH, and OSA.
\*p<0.05, \*\*p<0.01, \*\*\*p<0.005

In the sensitivity analysis, we controlled the maximum length of follow-up to ≤ 7.5 years and the results were consistent with the primary analysis. After adjusting for all the covariates in model 5 (in model 4 for VIM), patients in the fourth quartile of SD, CV, and VIM of BMI were significantly associated with higher HRs for incident HFpEF and HFrEF compared with those in the first quartile and heightened ASV of BMI was associated with increased risk of HFpEF.

## Discussion

We have performed a comprehensive analysis of the differential association between the long-term variability of BMI and the risk of HFrEF and HFpEF in a large cohort of patients with overweight or obesity from hospital settings. We found the larger intra-individual BMI variability, measured as a continuous variable or as quartiles, is associated with a greater risk of developing both HFrEF and HFpEF, after adjusting for all potential risk factors, including a wide array of comorbidities and mean BMI. More importantly, our results revealed consistent associations between all four BMI variability metrics and the risk of HFpEF. Additionally, we observed uniform findings for SD, CV, and VIM of BMI in relation to the risk of HFrEF. This emphasizes the importance of controlling weight variability in weight management strategies for overweight and obese patients to reduce the risk of HF. Furthermore, we validated that the baseline BMI is a risk factor for HF, and the association was more significant for HFpEF compared to HFrEF. This study highlights the potential feasibility and utility of using the different metrics to capture the variability of BMI derived from clinical data to predict the risk of HF in clinical practice settings. Our results also contribute to the growing body of evidence on reducing weight fluctuation in patients with overweight and obesity to mitigate the risk of adverse cardiovascular outcomes.

Our study provides unique and significant insights in several ways. 1.) We expanded the previous work by evaluating the heterogeneous association between BMI variability and HFpEF and HFrEF. Few studies have investigated the relationship between BMI fluctuation and incident HF in a cohort of obese and overweight patients. A previous study assessed the association of BMI variability with HF in overweight or obese patients with T2DM, but they were not able to identify HFrEF and HFpEF.^19^ The availability of image reports, as provided by the standardized performance of ECHO in hospital settings, allows us to accurately identify the specific subtype of incident HF cases. Our study addressed the current lack of evidence regarding the relationship between BMI variability and different subtypes of heart failure, thereby expanding the current knowledge in the field of obesity management and the etiology of HF. The positive association between BMI variability and both HFrEF and HFpEF in our study aligns with the findings of a previous study that also observed a positive association between BMI variability and overall heart failure.^19^ 2.) We used a large sample of patients with longitudinal follow-up in real-world hospital settings, as previous studies typically relied on data from randomized clinical trials or disease registries. It is widely recognized that findings derived from the clinical trial may not be always generalizable to real-world clinical settings. Clinical trials typically adhere to standardized weight measurement protocols, often involving double measurement from well-trained medical staff members, to ensure the accuracy and reliability of the data.^23,24^ However, it is important to recognize the challenges to follow strict patient eligibility and measurement protocols employed in clinical trials in real-world everyday clinical practice, where variations in measurement techniques and expertise among healthcare providers can occur.^25^ Nevertheless, using the clinical data derived from EHR, we capture the inherent complexities and variations in the routine clinical environment. Therefore, our results provide a broader and more universal understanding of the prognostic utility of BMI variability in assessing the risk of HF in real-world patient populations.

The exact mechanisms linking the variability in BMI to HF, especially for HFpEF, in patients with overweight or obesity still remain incompletely understood but are likely to be complex and multifactorial. Our hypothetical explanation is based on the distinct pathophysiology of HFpEF and HFrEF, considering the prevalence of comorbidities and the highly different responses of HFpEF and HFrEF to guideline-directed medical therapies. Given the well-described positive correlation between increasing obesity level with elevated risk of developing HFpEF, it is not surprising to observe a presumed relationship between higher variability in BMI and an increased risk of HFpEF development. One potential mechanism may involve the long-term implication of body weight fluctuation on the development and progression of metabolic syndrome, as patients with metabolic syndrome are known to have a substantially higher risk of developing HFpEF.^26–29^ Another potential pathway involves chronic systemic low-grade inflammation, as evidenced by higher concentrations of C-reactive protein in patients with large weight fluctuations. This chronic inflammation, in turn, could contribute to the development of HFpEF.^30^ Additionally, it is worth noting that the variability in BMI also predicts a higher risk of HFrEF, despite the previous study demonstrating a relatively weaker association between body weight and HFrEF compared to HFpEF.^31^ The increased weight cycle is associated with an increased risk of atrial fibrillation, which is a well-known risk factor for both HFrEF and HFpEF.^32^. Further research is necessary to investigate the underlying pathways linking body weight variability to the development of HFrEF and HFpEF.

The implications of our findings are meaningful for both clinicians and the general overweight or obese population. It is crucial to minimize body weight fluctuation when pursuing weight loss goals, as it has the potential to reduce the risk of HF and alleviate the considerable socioeconomic burden associated with this condition. It is particularly relevant for healthcare professionals who recommend dieting as a preventive strategy for obesity-related complications. Furthermore, the impact of our study can extend to the general population with obesity, as dieting is a subject of interest to many individuals. By emphasizing the importance of stable and consistent progress in weight loss, healthcare providers can help patients achieve their weight management goals while minimizing the risk of developing HF associated with significant weight fluctuations.

Our study possesses several strengths including 1). To our acknowledge, this analysis represents the first investigation that evaluates the differential association between variability of BMI with the incidence of subtypes of HF, facilitated by ECHO data stored in EHR allowing the precise identification of subjects with clinical HF with HFpEF and HFrEF. Recall that HF is a clinical syndrome and that having a normal LVEF alone does not identify HF. These subjects were identified from hospital EHR requiring a HF diagnosis. 2). Unlike most of the cohort studies where the BMI is measured only a few times, variability of BMI is captured from a high frequency of data over a long period from disparate clinical scenarios in our analysis. On average, each patient has a 19.69 BMI measurement over the 5-year BMI variability assessment period, which precisely and accurately reflects the BMI variability in the long term. 3). Compared to previous cohort studies investigating the relationship between BMI variability and cardiovascular outcomes, our cohort contains a substantially larger group of patients with incident HF, allowing us to generate more robust results. 4). We addressed the situation of malignancy tumors which is typically not considered in other studies. 5) Importantly, our study has a sample that is reflective of the US HF population (50-70% are female and 9-15% are black patients).

Our study should be interpreted in the context of a few limitations. 1.) Our study specifically focuses on classifying the HF by ejection fraction. However, we are not able to try other HF classifications, such as New York Heart Failure classification, acute vs chronic, ischemic vs non-ischemic, or primary myocardial disease vs secondary neurohormonal activity, due to the lack of data.^33^ 2.) Our study is a retrospective observational study, we could not draw any causal conclusion from our results. 3) Error in EHR is common, which could lead to underdiagnosis of certain comorbidities.

## Conclusion

In this study with a large group of overweight and obese patients, a higher degree of BMI variability was associated with an increased risk of developing HF compared to those with lower BMI variability. Our findings demonstrate that variability in BMI obtained from various clinical settings can serve as an accessible and valuable indicator for the risk of both HFrEF and HFpEF in real-world healthcare environments.

## Disclosure

It is important to acknowledge the potential conflict of interest among the contributors to this research. ZY, MZ, NF, and LW report receiving research funding from Eli Lilly and Company within the past 12 months. TPR, NCS, YC, and FS are employees of Eli Lilly and Company and own company stock while the research was being conducted.

## Author contribution

ZY played a vital role in the research process, data analysis, and composing the manuscript. YQ was involved in the study design, statistical method, and manuscript drafting. LW and YC were involved in the concept, data acquisition, study design, interpretation, comprehensive review and revision of the final manuscript, the final approval of the manuscripts, and project management. MZ and FN engaged in concept, study design, and discussion. TPR, NCS, and FS contributed to the study design, interpretation, discussion, review, and revision of the manuscript and final approval.

## Data Availability

All data produced in the present work are contained in the manuscript

## Reference

1. Center for Disease Control and Prevention. Heart Failure. Updated January 5, 2023. Accessed Feburary 23, 2023. https://www.cdc.gov/heartdisease/heart_failure.htm

2. Heart Failure Society of America. Heart Failure Facts & Information. Accessed Feburary 23, 2023. https://hfsa.org/patient-hub/heart-failure-facts-information

3. Adamczak DM, Oduah MT, Kiebalo T, et al. Heart Failure with Preserved Ejection Fraction-a Concise Review. Curr Cardiol Rep. 2020;22(9):82.

4. Borlaug BA, Paulus WJ. Heart failure with preserved ejection fraction: pathophysiology, diagnosis, and treatment. Eur Heart J. 2011;32(6):670–679.

5. Tsao CW, Aday AW, Almarzooq ZI, et al. Heart Disease and Stroke Statistics-2022 Update: A Report From the American Heart Association. Circulation. 2022;145(8):e153–e639.

6. Heidenreich PA, Bozkurt B, Aguilar D, et al. 2022 AHA/ACC/HFSA Guideline for the Management of Heart Failure: A Report of the American College of Cardiology/American Heart Association Joint Committee on Clinical Practice Guidelines. Circulation. 2022;145(18):e895–e1032.

7. Kenchaiah S, Evans JC, Levy D, et al. Obesity and the risk of heart failure. N Engl J Med. 2002;347(5):305–313.

8. Ho JE, Lyass A, Lee DS, et al. Predictors of new-onset heart failure: differences in preserved versus reduced ejection fraction. Circ Heart Fail. 2013;6(2):279–286.

9. Gong FF, Jelinek MV, Castro JM, et al. Risk factors for incident heart failure with preserved or reduced ejection fraction, and valvular heart failure, in a community-based cohort. Open Heart. 2018;5(2):e000782.

10. Huang P, Guo Z, Liang W, et al. Weight Change and Mortality Risk in Heart Failure With Preserved Ejection Fraction. Front Cardiovasc Med. 2021;8:681726.

11. Oga EA, Eseyin OR. The Obesity Paradox and Heart Failure: A Systematic Review of a Decade of Evidence. J Obes. 2016;2016:9040248.

12. Abebe TB, Gebreyohannes EA, Tefera YG, Abegaz TM. Patients with HFpEF and HFrEF have different clinical characteristics but similar prognosis: a retrospective cohort study. BMC Cardiovasc Disord. 2016;16(1):232.

13. Powell-Wiley TM, Poirier P, Burke LE, et al. Obesity and Cardiovascular Disease: A Scientific Statement From the American Heart Association. Circulation. 2021;143(21):e984–e1010.

14. Massey RJ, Siddiqui MK, Pearson ER, Dawed AY. Weight variability and cardiovascular outcomes: a systematic review and meta-analysis. Cardiovasc Diabetol. 2023;22(1):5.

15. Jeong S, Choi S, Chang J, et al. Association of weight fluctuation with cardiovascular disease risk among initially obese adults. Sci Rep. 2021;11(1):10152.

16. Li Y, Yu Y, Wu Y, et al. Association of Body-Weight Fluctuation With Outcomes in Heart Failure With Preserved Ejection Fraction. Front Cardiovasc Med. 2021;8:689591.

17. Kaze AD, Santhanam P, Erqou S, Ahima RS, Bertoni AG, Echouffo-Tcheugui JB. Body Weight Variability and Risk of Cardiovascular Outcomes and Death in the Context of Weight Loss Intervention Among Patients With Type 2 Diabetes. JAMA Netw Open. 2022;5(2):e220055.

18. Bangalore S, Fayyad R, Laskey R, DeMicco DA, Messerli FH, Waters DD. Body-Weight Fluctuations and Outcomes in Coronary Disease. N Engl J Med. 2017;376(14):1332–1340.

19. Kaze AD, Erqou S, Santhanam P, et al. Variability of adiposity indices and incident heart failure among adults with type 2 diabetes. Cardiovasc Diabetol. 2022;21(1):16.

20. Kwon S, Lee SR, Choi EK, et al. Visit-to-visit variability of metabolic parameters and risk of heart failure: A nationwide population-based study. Int J Cardiol. 2019;293:153–158.

21. Philipson H, Ekman I, Forslund HB, Swedberg K, Schaufelberger M. Salt and fluid restriction is effective in patients with chronic heart failure. Eur J Heart Fail. 2013;15(11):1304–1310.

22. Bozkurt B, Coats AJ, Tsutsui H, et al. Universal Definition and Classification of Heart Failure: A Report of the Heart Failure Society of America, Heart Failure Association of the European Society of Cardiology, Japanese Heart Failure Society and Writing Committee of the Universal Definition of Heart Failure. J Card Fail. 2021.

23. Ryan DH, Espeland MA, Foster GD, et al. Look AHEAD (Action for Health in Diabetes): design and methods for a clinical trial of weight loss for the prevention of cardiovascular disease in type 2 diabetes. Control Clin Trials. 2003;24(5):610–628.

24. Fitzgibbon ML, Stolley MR, Schiffer L, Sharp LK, Singh V, Dyer A. Obesity reduction black intervention trial (ORBIT): 18-month results. Obesity (Silver Spring). 2010;18(12):2317–2325.

25. Greenwood JL, Narus SP, Leiser J, Egger MJ. Measuring body mass index according to protocol: how are height and weight obtained? J Healthc Qual. 2011;33(3):28–36.

26. Purwowiyoto SL, Prawara AS. Metabolic syndrome and heart failure: mechanism and management. Med Pharm Rep. 2021;94(1):15–21.

27. Chin YR, So ES. The effects of weight fluctuation on the components of metabolic syndrome: a 16-year prospective cohort study in South Korea. Arch Public Health. 2021;79(1):21.

28. Vergnaud AC, Bertrais S, Oppert JM, et al. Weight fluctuations and risk for metabolic syndrome in an adult cohort. Int J Obes (Lond). 2008;32(2):315–321.

29. Zhang H, Tamakoshi K, Yatsuya H, et al. Long-term body weight fluctuation is associated with metabolic syndrome independent of current body mass index among Japanese men. Circ J. 2005;69(1):13–18.

30. Tamakoshi K, Yatsuya H, Kondo T, et al. Long-term body weight variability is associated with elevated C-reactive protein independent of current body mass index among Japanese men. Int J Obes Relat Metab Disord. 2003;27(9):1059–1065.

31. Savji N, Meijers WC, Bartz TM, et al. The Association of Obesity and Cardiometabolic Traits With Incident HFpEF and HFrEF. JACC Heart Fail. 2018;6(8):701–709.

32. Lee HJ, Choi EK, Han KD, et al. Bodyweight fluctuation is associated with increased risk of incident atrial fibrillation. Heart Rhythm. 2020;17(3):365–371.

33. Banerjee A, Dashtban A, Chen S, et al. Identifying subtypes of heart failure from three electronic health record sources with machine learning: an external, prognostic, and genetic validation study. Lancet Digit Health. 2023;5(6):e370–e379.

