## Supplemental files for "Body Mass Index (BMI) and BMI Variability are Risk Factors for Heart Failure with Preserved and Reduced Ejection Fraction in a Longitudinal Cohort Study Using Real-World Electronic Health Records"

**Table S1. Comparison of Baseline Demographics and Characteristics of Study Patients by Baseline BMI Categories**

**Table S2. Comparison of Baseline Demographics and Characteristics of Study Patients by HF Subtypes**

**Table S3. The Association between Clinical Characteristics with Incident Heart Failure by Quartiles of SD of BMI**

**Table S4. The Association between Clinical Characteristics with Incident Heart Failure by Quartiles of CV of BMI**

**Table S5. The Association between Clinical Characteristics with Incident Heart Failure by Quartiles of VIM of BMI**

**Table S6. The Association between Clinical Characteristics with Incident Heart Failure by Quartiles of ASV of BMI**

**Table S7. The Association between BMI Variability with Heart Failure by Quartiles of BMI Variability with Maximum of 7.5 Years of Follow-up**

**Table S1. Comparison of Baseline Demographics and Characteristics of Study Patients by Baseline BMI Categories**

| **BMI Categories** |  | **Study Patients** | **<27** | **27-30** | **Obesity Class I** | **Obesity Class II** | **Obesity Class III** | **P-Value** |
| --- | --- | --- | --- | --- | --- | --- | --- | --- |
| **n** |  | 51444 | 2205 | 10971 | 14454 | 12250 | 11564 |  |
| **Age, mean (SD)** |  | 52.83 (14.66) | 57.52 (15.95) | 56.25 (14.47) | 54.17 (14.54) | 52.09 (13.94) | 47.80 (13.99) | <0.001 |
| **Gender, n (%)** | FEMALE | 31408 (61.05) | 1335 (60.54) | 5748 (52.39) | 8175 (56.56) | 7764 (63.38) | 8386 (72.52) | <0.001 |
|  | MALE | 20036 (38.95) | 870 (39.46) | 5223 (47.61) | 6279 (43.44) | 4486 (36.62) | 3178 (27.48) |  |
| **Race, n (%)** | WHITE | 44521 (86.54) | 1966 (89.16) | 9868 (89.95) | 12698 (87.85) | 10459 (85.38) | 9530 (82.41) | <0.001 |
|  | BLACK | 6257 (12.16) | 206 (9.34) | 897 (8.18) | 1557 (10.77) | 1673 (13.66) | 1924 (16.64) |  |
|  | OTHER | 666 (1.29) | 33 (1.50) | 206 (1.88) | 199 (1.38) | 118 (0.96) | 110 (0.95) |  |
| **Smoking Status, mean (SD)** |  | 6610 (12.85) | 319 (14.47) | 1344 (12.25) | 1821 (12.60) | 1583 (12.92) | 1543 (13.34) | 0.017 |
| **Num of BMI Records, mean (SD)** |  | 19.69 (10.86) | 22.13 (11.97) | 18.17 (9.72) | 19.18 (10.55) | 20.18 (11.21) | 20.79 (11.40) | <0.001 |
| **ASCVD, n (%)** |  | 10540 (20.49) | 561 (25.44) | 2385 (21.74) | 3168 (21.92) | 2523 (20.60) | 1903 (16.46) | <0.001 |
| **T2DM, n (%)** |  | 15850 (30.81) | 537 (24.35) | 2285 (20.83) | 4016 (27.78) | 4430 (36.16) | 4582 (39.62) | <0.001 |
| **HTN, n (%)** |  | 36908 (71.74) | 1454 (65.94) | 7177 (65.42) | 10185 (70.46) | 9261 (75.60) | 8831 (76.37) | <0.001 |
| **HLP, n (%)** |  | 36862 (71.65) | 1569 (71.16) | 8078 (73.63) | 10606 (73.38) | 8927 (72.87) | 7682 (66.43) | <0.001 |
| **CKD, n (%)** |  | 4373 (8.50) | 219 (9.93) | 860 (7.84) | 1252 (8.66) | 1077 (8.79) | 965 (8.34) | 0.007 |
| **NAFLD/NASH, n (%)** |  | 4289 (8.34) | 139 (6.30) | 536 (4.89) | 1071 (7.41) | 1210 (9.88) | 1333 (11.53) | <0.001 |
| **OSA, n (%)** |  | 11248 (21.86) | 232 (10.52) | 1192 (10.87) | 2536 (17.55) | 3174 (25.91) | 4114 (35.58) | <0.001 |
| **Mean HR, mean (SD)** |  | 77.09 (8.57) | 75.56 (8.44) | 75.03 (8.29) | 76.42 (8.46) | 77.67 (8.42) | 79.59 (8.47) | <0.001 |
| **Mean SBP, mean (SD)** |  | 129.21 (9.40) | 127.06 (9.94) | 127.91 (9.76) | 128.80 (9.38) | 129.90 (9.15) | 130.65 (8.95) | <0.001 |
| **Mean DBP, mean (SD)** |  | 78.64 (5.75) | 76.30 (5.64) | 77.52 (5.75) | 78.36 (5.70) | 79.24 (5.62) | 79.87 (5.61) | <0.001 |
| **SD of HR, mean (SD)** |  | 9.16 (3.43) | 9.18 (3.46) | 8.78 (3.39) | 9.06 (3.41) | 9.17 (3.39) | 9.62 (3.47) | <0.001 |
| **SD of SBP, mean (SD)** |  | 11.95 (3.86) | 12.13 (3.90) | 11.77 (4.01) | 11.90 (3.84) | 11.96 (3.79) | 12.15 (3.81) | <0.001 |
| **SD of DBP, mean (SD)** |  | 7.74 (2.14) | 7.68 (2.02) | 7.54 (2.05) | 7.67 (2.09) | 7.74 (2.13) | 8.02 (2.29) | <0.001 |
| **Mean BMI, mean (SD)** |  | 35.30 (6.45) | 27.93 (1.43) | 28.75 (1.35) | 32.39 (2.17) | 37.18 (2.11) | 44.57 (4.59) | <0.001 |
| **Delta BMI, mean (SD)** |  | 0.23 (2.75) | -2.55 (2.01) | -0.24 (1.63) | 0.05 (2.24) | 0.24 (2.46) | 1.42(3.81) | <0.001 |
| **Quartile of CV of BMI** | Quartile1 | 12861 (25.00) | 332 (15.06) | 3763 (34.30) | 3673 (25.41) | 3047 (24.87) | 2046 (17.69) | <0.001 |
|  | Quartile2 | 12861 (25.00) | 447 (20.27) | 3021 (27.54) | 3752 (25.96) | 3114 (25.42) | 2527 (21.85) |  |
|  | Quartile3 | 12861 (25.00) | 607 (27.53) | 2383 (21.72) | 3683 (25.48) | 3174 (25.91) | 3014 (26.06) |  |
|  | Quartile4 | 12861 (25.00) | 819 (37.14) | 1804 (16.44) | 3346 (23.15) | 2915 (23.80) | 3977 (34.39) |  |

Abbreviations: BMI, body mass index; HF, heart failure; HFpEF, heart failure with preserved ejection fraction; HFrEF, heart failure with rejected ejection fraction; SD, standard deviation; CV, coefficient of variation; VIM, variability independent of mean; ASV, average successive variability; ASCVD, atherosclerotic cardiovascular disease; T2DM, type 2 diabetes mellitus; HTN, hypertension; HLP, hyperlipidemia; CKD, chronic kidney disease; NAFLD/NASH, nonalcoholic fatty liver disease/nonalcoholic steatohepatitis; OSA, obstructive sleep apnea; SBP, systolic blood pressure; DBP, diastolic blood pressure; HR, heart rate.

**Table S2 Comparison of Baseline Demographics and Characteristics of Study Patients by HF Subtypes**

| **HF Type** |  | **Study Patients** | **No HF** | **HFpEF** | **HFrEF** | **P-Value** |
| --- | --- | --- | --- | --- | --- | --- |
| **n** |  | 51444 | 48555 | 1871 | 1018 |  |
| **Age, mean (SD)** |  | 52.83 (14.66) | 52.14 (14.52) | 64.42 (11.93) | 64.35 (11.91) | <0.001 |
| **GENDER, n (%)** | FEMALE | 31408 (61.05) | 29845 (61.47) | 1129 (60.34) | 434 (42.63) | <0.001 |
|  | MALE | 20036 (38.95) | 18710 (38.53) | 742 (39.66) | 584 (57.37) |  |
| **RACE, n (%)** | WHITE | 44521 (86.54) | 42019 (86.54) | 1620 (86.58) | 882 (86.64) | 0.990 |
|  | BLACK | 6257 (12.16) | 5906 (12.16) | 229 (12.24) | 122 (11.98) |  |
|  | OTHER | 666 (1.29) | 630 (1.30) | 22 (1.18) | 14 (1.38) |  |
| **Smoking Status, mean (SD)** |  | 6610 (12.85) | 6255 (12.88) | 222 (11.87) | 133 (13.06) | 0.426 |
| **Num of BMI Records, mean (SD)** |  | 19.69 (10.86) | 19.53 (10.76) | 23.34 (12.67) | 20.73 (10.56) | <0.001 |
| **Baseline BMI Categories, n (%)** | <27 | 2205 (4.29) | 2061 (4.24) | 87 (4.65) | 57 (5.60) | <0.001 |
|  | Overweight | 10971 (21.33) | 10449 (21.52) | 285 (15.23) | 237 (23.28) |  |
|  | Obesity I | 14454 (28.10) | 13697 (28.21) | 484 (25.87) | 273 (26.82) |  |
|  | Obesity II | 12250 (23.81) | 11555 (23.80) | 458 (24.48) | 237 (23.28) |  |
|  | Obesity III | 11564 (22.48) | 10793 (22.23) | 557 (29.77) | 214 (21.02) |  |
| **ASCVD, n (%)** |  | 10540 (20.49) | 9125 (18.79) | 861 (46.02) | 554 (54.42) | <0.001 |
| **T2DM, n (%)** |  | 15850 (30.81) | 14348 (29.55) | 946 (50.56) | 556 (54.62) | <0.001 |
| **HTN, n (%)** |  | 36908 (71.74) | 34204 (70.44) | 1753 (93.69) | 951 (93.42) | <0.001 |
| **HLP, n (%)** |  | 36862 (71.65) | 34336 (70.72) | 1628 (87.01) | 898 (88.21) | <0.001 |
| **CKD, n (%)** |  | 4373 (8.50) | 3728 (7.68) | 437 (23.36) | 208 (20.43) | <0.001 |
| **NAFLD/NASH, n (%)** |  | 4289 (8.34) | 4020 (8.28) | 193 (10.32) | 76 (7.47) | 0.005 |
| **OSA, n (%)** |  | 11248 (21.86) | 10378 (21.37) | 608 (32.50) | 262 (25.74) | <0.001 |
| **Mean HR, mean (SD)** |  | 77.09 (8.57) | 77.22 (8.56) | 75.10 (8.62) | 74.81 (8.45) | <0.001 |
| **Mean SBP, mean (SD)** |  | 129.21 (9.40) | 128.98 (9.32) | 133.49 (9.69) | 132.30 (9.97) | <0.001 |
| **Mean DBP, mean (SD)** |  | 78.64 (5.75) | 78.75 (5.70) | 76.67 (6.09) | 77.23 (6.58) | <0.001 |
| **SD of HR, mean (SD)** |  | 9.16 (3.43) | 9.16 (3.43) | 9.20 (3.26) | 9.09 (3.49) | 0.689 |
| **SD of SBP, mean (SD)** |  | 11.95 (3.86) | 11.83 (3.81) | 14.19 (4.18) | 13.62 (4.17) | <0.001 |
| **SD of DBP, mean (SD)** |  | 7.74 (2.14) | 7.71 (2.13) | 8.34 (2.24) | 8.18 (2.22) | <0.001 |
| **Mean BMI, mean (SD)** |  | 35.30 (6.45) | 35.25 (6.44) | 36.69 (6.74) | 35.16 (6.38) | <0.001 |
| **Delta BMI, mean (SD)** |  | 0.23 (2.75) | 0.25 (2.74) | -0.09 (2.96) | -0.39 (2.74) | <0.001 |

Abbreviations: BMI, body mass index; HF, heart failure; HFpEF, heart failure with preserved ejection fraction; HFrEF, heart failure with rejected ejection fraction; SD, standard deviation; CV, coefficient of variation; VIM, variability independent of mean; ASV, average successive variability; ASCVD, atherosclerotic cardiovascular disease; T2DM, type 2 diabetes mellitus; HTN, hypertension; HLP, hyperlipidemia; CKD, chronic kidney disease; NAFLD/NASH, nonalcoholic fatty liver disease/nonalcoholic steatohepatitis; OSA, obstructive sleep apnea; SBP, systolic blood pressure; DBP, diastolic blood pressure; HR, heart rate.

**Table S3. The Association between Clinical Characteristics with Incident Heart Failure by Quartiles of SD of BMI**

| **Risk Factors** |  | **Models** | | | | | | | | | |
| --- | --- | --- | --- | --- | --- | --- | --- | --- | --- | --- | --- |
|  |  | **Model1** | | **Model2** | | **Model3** | | **Model4** | | **Model5** | |
| **HF Type** |  | HFpEF | HFrEF | HFpEF | HFrEF | HFpEF | HFrEF | HFpEF | HFrEF | HFpEF | HFrEF |
| **Age** |  | 1.08 (1.08-1.09)*** | 1.08 (1.08-1.09)*** | 1.09 (1.09-1.10)*** | 1.09 (1.08-1.09)*** | 1.08 (1.07-1.08)*** | 1.07 (1.06-1.08)*** | 1.08 (1.07-1.08)*** | 1.07 (1.06-1.08)*** | 1.08 (1.07-1.08)*** | 1.07 (1.06-1.08)*** |
| **Gender** | Female | Reference | Reference | Reference | Reference | Reference | Reference | Reference | Reference | Reference | Reference |
|  | Male | 1.19 (1.08-1.31)*** | 2.35 (2.07-2.67)*** | 1.29 (1.17-1.42)*** | 2.44 (2.14-2.77)*** | 1.04 (0.95-1.15) | 1.92 (1.68-2.19)*** | 1.06 (0.96-1.17) | 1.95 (1.70-2.22)*** | 1.08 (0.97-1.19) | 1.97 (1.73-2.25)*** |
| **Race** | White | Reference | Reference | Reference | Reference | Reference | Reference | Reference | Reference | Reference | Reference |
|  | Black | 1.14 (0.99-1.31) | 1.34 (1.11-1.63)*** | 1.09 (0.95-1.26) | 1.32 (1.08-1.60)*** | 1.00 (0.86-1.15) | 1.15 (0.95-1.41) | 0.95 (0.82-1.10) | 1.12 (0.92-1.37) | 0.94 (0.81-1.08) | 1.11 (0.91-1.35) |
|  | Other | 1.13 (0.74-1.72) | 1.41 (0.83-2.39) | 1.21 (0.80-1.85) | 1.45 (0.86-2.47) | 1.25 (0.82-1.91) | 1.52 (0.90-2.59) | 1.25 (0.82-1.90) | 1.53 (0.90-2.59) | 1.27 (0.83-1.93) | 1.54 (0.91-2.62) |
| **Smoking status** |  | 1.50 (1.30-1.74)*** | 1.69 (1.40-2.04)*** | 1.64 (1.42-1.90)*** | 1.75 (1.45-2.11)*** | 1.59 (1.37-1.84)*** | 1.62 (1.34-1.96)*** | 1.56 (1.35-1.80)*** | 1.60 (1.32-1.93)*** | 1.56 (1.35-1.80)*** | 1.60 (1.32-1.93)*** |
| **Num of BMI records** |  | 1.03 (1.03-1.03)*** | 1.02 (1.01-1.02)*** | 1.03 (1.03-1.03)*** | 1.02 (1.01-1.02)*** | 1.02 (1.02-1.03)*** | 1.01 (1.00-1.01) | 1.02 (1.02-1.02)*** | 1.01 (1.00-1.01) | 1.02 (1.02-1.03)*** | 1.01 (1.00-1.01) |
| **BMI<27** |  | Reference | Reference | Reference | Reference | Reference | Reference | Reference | Reference | Reference | Reference |
| **Overweight** |  | Na | Na | 0.88 (0.69-1.12) | 1.01 (0.75-1.35) | 0.87 (0.68-1.10) | 0.99 (0.74-1.32) | 0.85 (0.67-1.08) | 0.98 (0.74-1.32) | 0.82 (0.64-1.04) | 0.95 (0.71-1.27) |
| **Obesity Class I** |  | Na | Na | 1.27 (1.01-1.60)* | 1.00 (0.75-1.34) | 1.17 (0.93-1.47) | 0.92 (0.69-1.22) | 1.18 (0.94-1.49) | 0.92 (0.69-1.23) | 1.00 (0.79-1.27) | 0.79 (0.58-1.07) |
| **Obesity Class II** |  | Na | Na | 1.66 (1.31-2.09)*** | 1.24 (0.93-1.66) | 1.40 (1.11-1.77)*** | 1.05 (0.78-1.41) | 1.44 (1.14-1.82)*** | 1.07 (0.80-1.44) | 1.04 (0.79-1.36) | 0.79 (0.55-1.14) |
| **Obesity Class III** |  | Na | Na | 2.83 (2.24-3.57)*** | 1.62 (1.20-2.20)*** | 2.26 (1.78-2.87)*** | 1.33 (0.98-1.81) | 2.33 (1.84-2.96)*** | 1.35 (0.99-1.84) | 1.30 (0.92-1.82) | 0.80 (0.49-1.30) |
| **ASCVD** |  | Na | Na | Na | Na | 1.76 (1.59-1.94)*** | 2.37 (2.08-2.71)*** | 1.70 (1.54-1.88)*** | 2.34 (2.04-2.67)*** | 1.71 (1.55-1.89)*** | 2.34 (2.05-2.68)*** |
| **T2DM** |  | Na | Na | Na | Na | 1.26 (1.14-1.38)*** | 1.66 (1.46-1.90)*** | 1.26 (1.14-1.39)*** | 1.66 (1.46-1.89)*** | 1.23 (1.12-1.36)*** | 1.63 (1.43-1.86)*** |
| **HTN** |  | Na | Na | Na | Na | 1.78 (1.46-2.16)*** | 1.72 (1.33-2.24)*** | 1.62 (1.33-1.97)*** | 1.63 (1.25-2.12)*** | 1.59 (1.31-1.94)*** | 1.61 (1.24-2.09)*** |
| **HLP** |  | Na | Na | Na | Na | 0.91 (0.79-1.06) | 0.90 (0.74-1.11) | 0.92 (0.80-1.07) | 0.92 (0.75-1.12) | 0.93 (0.80-1.07) | 0.92 (0.75-1.12) |
| **CKD** |  | Na | Na | Na | Na | 1.56 (1.39-1.75)*** | 1.34 (1.14-1.58)*** | 1.52 (1.35-1.70)*** | 1.31 (1.12-1.54)*** | 1.51 (1.35-1.69)*** | 1.31 (1.12-1.54)*** |
| **NAFLD/NASH** |  | Na | Na | Na | Na | 1.18 (1.01-1.37)* | 0.94 (0.74-1.19) | 1.17 (1.01-1.37)* | 0.93 (0.74-1.19) | 1.17 (1.00-1.37)* | 0.93 (0.73-1.18) |
| **OSA** |  | Na | Na | Na | Na | 1.31 (1.17-1.45)*** | 0.99 (0.85-1.15) | 1.32 (1.18-1.46)*** | 0.99 (0.85-1.15) | 1.28 (1.15-1.43)*** | 0.97 (0.83-1.13)*** |
| **SD of systolic BP** |  | Na | Na | Na | Na | Na | Na | 1.04 (1.04-1.05)*** | 1.03 (1.02-1.04)*** | 1.04 (1.04-1.05)*** | 1.03 (1.02-1.04)*** |
| **SD of HR** |  | Na | Na | Na | Na | Na | Na | 1.02 (1.01-1.04)*** | 1.03 (1.01-1.05)*** | 1.02 (1.01-1.04)*** | 1.03 (1.01-1.05)*** |
| **Mean BMI** |  | Na | Na | Na | Na | Na | Na | Na | Na | 1.04 (1.02-1.06)*** | 1.03 (1.01-1.06)** |
| **Quartile of SD of BMI** | Quartile2 | 1.36 (1.18-1.57)*** | 1.21 (1.01-1.45)* | 1.20 (1.04-1.39)* | 1.16 (0.97-1.39) | 1.17 (1.01-1.35)* | 1.14 (0.95-1.37) | 1.13 (0.98-1.31) | 1.12 (0.93-1.34) | 1.12 (0.97-1.30) | 1.11 (0.92-1.33) |
|  | Quartile3 | 1.76 (1.53-2.02)*** | 1.64 (1.38-1.96)*** | 1.37 (1.19-1.58)*** | 1.50 (1.24-1.80)*** | 1.29 (1.12-1.50)*** | 1.42 (1.18-1.71)*** | 1.22 (1.06-1.41)*** | 1.36 (1.13-1.64)*** | 1.20 (1.04-1.38)*** | 1.34 (1.11-1.61)*** |
|  | Quartile4 | 2.55 (2.22-2.92)*** | 2.06 (1.71-2.47)*** | 1.72 (1.48-1.99)*** | 1.74 (1.43-2.12)*** | 1.59 (1.37-1.84)*** | 1.63 (1.34-1.98)*** | 1.43 (1.23-1.66)*** | 1.51 (1.24-1.84)*** | 1.35 (1.16-1.57)*** | 1.43 (1.17-1.75)*** |

Abbreviations: BMI, body mass index; HF, heart failure; HFpEF, heart failure with preserved ejection fraction; HFrEF, heart failure with rejected ejection fraction; SD, standard deviation; CV, coefficient of variation; VIM, variability independent of mean; ASV, average successive variability; ASCVD, atherosclerotic cardiovascular disease; T2DM, type 2 diabetes mellitus; HTN, hypertension; HLP, hyperlipidemia; CKD, chronic kidney disease; NAFLD/NASH, nonalcoholic fatty liver disease/nonalcoholic steatohepatitis; OSA, obstructive sleep apnea; SBP, systolic blood pressure; HR, heart rate.

Model 1 adjusted for age, gender, race, smoking status, and the number of BMI Records.

Model 2 adjusted for variables in model 1 plus baseline BMI classes.

Model 3: adjusted for variables in model 2 plus ASCVD, T2DM, HTN, HLP, CKD, NAFLD/NASH, and OSA.

Model 4: adjusted for variables in model 3 plus SD of SBP and SD of HR.

Model 5: adjusted for variables in model 3 plus mean BMI.

*p<0.05, **p<0.01, ***p<0.005

**Table S4. The Association between Clinical Characteristics with Incident Heart Failure by Quartiles of CV of BMI**

| **Risk Factors** |  | **Models** | | | | | | | | | |
| --- | --- | --- | --- | --- | --- | --- | --- | --- | --- | --- | --- |
|  |  | **Model1** | | **Model2** | | **Model3** | | **Model4** | | **Model5** | |
| **HF Type** |  | **HFpEF** | **HFrEF** | **HFpEF** | **HFrEF** | **HFpEF** | **HFrEF** | **HFpEF** | **HFrEF** | **HFpEF** | **HFrEF** |
| **Age** |  | 1.08 (1.08-1.08)*** | 1.08 (1.07-1.09)*** | 1.09 (1.09-1.10)*** | 1.09 (1.08-1.09)*** | 1.08 (1.07-1.08)*** | 1.07 (1.06-1.08)*** | 1.08 (1.07-1.08)*** | 1.07 (1.06-1.08)*** | 1.08 (1.07-1.08)*** | 1.07 (1.06-1.08)*** |
| **Gender** | Female | Reference | Reference | Reference | Reference | Reference | Reference | Reference | Reference | Reference | Reference |
|  | Male | 1.13 (1.03-1.24)** | 2.27 (2.00-2.58)*** | 1.28 (1.16-1.41)*** | 2.42 (2.13-2.75)*** | 1.04 (0.94-1.14) | 1.91 (1.67-2.18)*** | 1.05 (0.95-1.16) | 1.94 (1.69-2.21)*** | 1.08 (0.97-1.19) | 1.97 (1.72-2.25)*** |
| **Race** | White | Reference | Reference | Reference | Reference | Reference | Reference | Reference | Reference | Reference | Reference |
|  | Black | 1.16 (1.00-1.33)* | 1.36 (1.12-1.66)*** | 1.09 (0.95-1.26) | 1.32 (1.09-1.61)** | 1.00 (0.86-1.15) | 1.16 (0.95-1.41) | 0.95 (0.82-1.10) | 1.12 (0.92-1.37) | 0.93 (0.81-1.08) | 1.11 (0.91-1.36) |
|  | Other | 1.13 (0.74-1.72) | 1.41 (0.83-2.39) | 1.22 (0.80-1.86) | 1.47 (0.86-2.49) | 1.26 (0.83-1.92) | 1.54 (0.91-2.61) | 1.25 (0.82-1.91) | 1.54 (0.91-2.61) | 1.27 (0.83-1.94) | 1.56 (0.92-2.64) |
| **Smoking status** |  | 1.51 (1.31-1.74)*** | 1.69 (1.40-2.04)*** | 1.65 (1.43-1.91)*** | 1.76 (1.46-2.12)*** | 1.60 (1.38-1.85)*** | 1.63 (1.35-1.97)*** | 1.56 (1.35-1.81)*** | 1.61 (1.33-1.94)*** | 1.56 (1.35-1.81)*** | 1.60 (1.33-1.94)*** |
| **Num of BMI records** |  | 1.03 (1.03-1.03)*** | 1.02 (1.01-1.02)*** | 1.03 (1.03-1.03)*** | 1.02 (1.01-1.02)*** | 1.02 (1.02-1.03)*** | 1.01 (1.00-1.01)* | 1.02 (1.02-1.02)*** | 1.01 (1.00-1.01) | 1.02 (1.02-1.03)*** | 1.01 (1.00-1.01) |
| **BMI<27** |  | Reference | Reference | Reference | Reference | Reference | Reference | Reference | Reference | Reference | Reference |
| **Overweight** |  | Na | Na | 0.88 (0.69-1.12) | 1.01 (0.75-1.35) | 0.86 (0.68-1.10) | 0.99 (0.74-1.32) | 0.85 (0.67-1.08) | 0.98 (0.73-1.32) | 0.81 (0.64-1.04) | 0.94 (0.70-1.26) |
| **Obesity Class I** |  | Na | Na | 1.33 (1.06-1.68)* | 1.06 (0.79-1.41) | 1.22 (0.97-1.53) | 0.96 (0.72-1.27) | 1.22 (0.97-1.53) | 0.95 (0.72-1.27) | 1.01 (0.79-1.28) | 0.80 (0.59-1.08) |
| **Obesity Class II** |  | Na | Na | 1.83 (1.45-2.31)*** | 1.39 (1.03-1.86)* | 1.52 (1.20-1.92)*** | 1.15 (0.86-1.55) | 1.53 (1.21-1.94)*** | 1.16 (0.86-1.56) | 1.05 (0.80-1.38) | 0.81 (0.57-1.17) |
| **Obesity Class III** |  | Na | Na | 3.32 (2.63-4.18)*** | 1.93 (1.43-2.61)*** | 2.58 (2.04-3.27)*** | 1.54 (1.13-2.09)*** | 2.58 (2.04-3.26)*** | 1.53 (1.13-2.08)*** | 1.33 (0.95-1.86)*** | 0.82 (0.50-1.33)*** |
| **ASCVD** |  | Na | Na | Na | Na | 1.75 (1.59-1.93)*** | 2.37 (2.08-2.71)*** | 1.70 (1.54-1.88)*** | 2.34 (2.04-2.67)*** | 1.71 (1.55-1.89)*** | 2.35 (2.05-2.68)*** |
| **T2DM** |  | Na | Na | Na | Na | 1.26 (1.15-1.39)*** | 1.67 (1.46-1.90)*** | 1.27 (1.15-1.40)*** | 1.67 (1.46-1.90)*** | 1.23 (1.12-1.36)*** | 1.63 (1.43-1.86)*** |
| **HTN** |  | Na | Na | Na | Na | 1.79 (1.47-2.17)*** | 1.73 (1.34-2.25)*** | 1.62 (1.33-1.98)*** | 1.64 (1.26-2.12)*** | 1.59 (1.31-1.94)*** | 1.61 (1.24-2.09)*** |
| **HLP** |  | Na | Na | Na | Na | 0.91 (0.79-1.05) | 0.90 (0.74-1.10) | 0.92 (0.80-1.06) | 0.91 (0.75-1.12) | 0.93 (0.80-1.07) | 0.92 (0.75-1.12) |
| **CKD** |  | Na | Na | Na | Na | 1.56 (1.40-1.75)*** | 1.35 (1.15-1.58)*** | 1.52 (1.35-1.70)*** | 1.32 (1.12-1.55)*** | 1.51 (1.35-1.69)*** | 1.31 (1.12-1.54)*** |
| **NAFLD/NASH** |  | Na | Na | Na | Na | 1.18 (1.01-1.38)* | 0.94 (0.74-1.20) | 1.18 (1.01-1.37)* | 0.94 (0.74-1.19) | 1.17 (1.01-1.37)* | 0.93 (0.74-1.19) |
| **OSA** |  | Na | Na | Na | Na | 1.31 (1.18-1.46) | 0.99 (0.85-1.15) | 1.32 (1.19-1.47)*** | 0.99 (0.85-1.16) | 1.28 (1.15-1.43)*** | 0.97 (0.83-1.13) |
| **SD of systolic BP** |  | Na | Na | Na | Na | Na | Na | 1.04 (1.04-1.05)*** | 1.03 (1.02-1.04)*** | 1.05 (1.04-1.05)*** | 1.03 (1.02-1.04)*** |
| **SD of HR** |  | Na | Na | Na | Na | Na | Na | 1.03 (1.01-1.04)*** | 1.03 (1.01-1.05)*** | 1.02 (1.01-1.04)*** | 1.03 (1.01-1.05)*** |
| **Mean BMI** |  | Na | Na | Na | Na | Na | Na | Na | Na | 1.04 (1.03-1.06)*** | 1.04 (1.02-1.06) |
| **Quartile of CV of BMI** | Quartile2 | 1.12 (0.97-1.28) | 1.29 (1.08-1.54)** | 1.08 (0.94-1.23) | 1.27 (1.06-1.51)** | 1.05 (0.92-1.21) | 1.24 (1.04-1.49)* | 1.03 (0.90-1.18) | 1.22 (1.02-1.46)* | 1.03 (0.90-1.18) | 1.22 (1.02-1.46)* |
|  | Quartile3 | 1.26 (1.10-1.44)*** | 1.45 (1.21-1.73)*** | 1.18 (1.03-1.35)* | 1.40 (1.17-1.67)*** | 1.13 (0.99-1.30) | 1.35 (1.13-1.62)*** | 1.08 (0.94-1.24) | 1.30 (1.09-1.56)*** | 1.07 (0.94-1.23) | 1.29 (1.08-1.55)*** |
|  | Quartile4 | 1.66 (1.45-1.89)*** | 1.73 (1.44-2.07)*** | 1.49 (1.30-1.70)*** | 1.63 (1.35-1.96)*** | 1.39 (1.22-1.59)*** | 1.54 (1.28-1.85)*** | 1.27 (1.11-1.45)*** | 1.44 (1.19-1.74)*** | 1.25 (1.09-1.43)*** | 1.41 (1.17-1.70)*** |

Abbreviations: BMI, body mass index; HF, heart failure; HFpEF, heart failure with preserved ejection fraction; HFrEF, heart failure with rejected ejection fraction; SD, standard deviation; CV, coefficient of variation; VIM, variability independent of mean; ASV, average successive variability; ASCVD, atherosclerotic cardiovascular disease; T2DM, type 2 diabetes mellitus; HTN, hypertension; HLP, hyperlipidemia; CKD, chronic kidney disease; NAFLD/NASH, nonalcoholic fatty liver disease/nonalcoholic steatohepatitis; OSA, obstructive sleep apnea; SBP, systolic blood pressure; HR, heart rate.

Model 1 adjusted for age, gender, race, smoking status, and the number of BMI Records.

Model 2 adjusted for variables in model 1 plus baseline BMI classes.

Model 3: adjusted for variables in model 2 plus ASCVD, T2DM, HTN, HLP, CKD, NAFLD/NASH, and OSA.

Model 4: adjusted for variables in model 3 plus SD of SBP and SD of HR.

Model 5: adjusted for variables in model 4 plus mean BMI.

*p<0.05, **p<0.01, ***p<0.005

**Table S5. The Association between Clinical Characteristics with Incident Heart Failure by Quartiles of VIM of BMI**

| **Risk Factors** |  | **Models** | | | | | | | | | |
| --- | --- | --- | --- | --- | --- | --- | --- | --- | --- | --- | --- |
|  |  | **Model1** | | **Model2** | | **Model3** | | **Model4** | | **Model5** | |
| **HF Type** |  | HFpEF | HFrEF | HFpEF | HFrEF | HFpEF | HFrEF | HFpEF | HFrEF | HFpEF | HFrEF |
| **Age** |  | 1.08 (1.07-1.08)*** | 1.08 (1.07-1.09)*** | 1.09 (1.09-1.10)*** | 1.09 (1.08-1.09)*** | 1.08 (1.07-1.08)*** | 1.07 (1.06-1.08)*** | 1.08 (1.07-1.08)*** | 1.07 (1.06-1.08)*** | Na | Na |
| **Gender** | Female | Reference | Reference | Reference | Reference | Reference | Reference | Reference | Reference | Na | Na |
|  | Male | 1.11 (1.01-1.22)* | 2.24 (1.97-2.54)*** | 1.28 (1.16-1.41)*** | 2.41 (2.12-2.74)*** | 1.04 (0.94-1.14) | 1.90 (1.66-2.17)*** | 1.05 (0.95-1.16) | 1.93 (1.69-2.20)*** | Na | Na |
| **Race** | White | Reference | Reference | Reference | Reference | Reference | Reference | Reference | Reference | Na | Na |
|  | Black | 1.16 (1.01-1.34)* | 1.37 (1.12-1.66)*** | 1.09 (0.95-1.26) | 1.32 (1.09-1.60)** | 1.00 (0.86-1.15) | 1.16 (0.95-1.41) | 0.95 (0.82-1.10) | 1.12 (0.92-1.37) | Na | Na |
|  | Other | 1.13 (0.74-1.73) | 1.40 (0.83-2.38) | 1.22 (0.80-1.86) | 1.46 (0.86-2.48) | 1.26 (0.83-1.92) | 1.53 (0.90-2.60) | 1.25 (0.82-1.91) | 1.53 (0.90-2.60) | Na | Na |
| **Smoking status** |  | 1.52 (1.31-1.75)*** | 1.70 (1.41-2.05)*** | 1.65 (1.43-1.91)*** | 1.77 (1.46-2.13)*** | 1.59 (1.38-1.84)*** | 1.63 (1.35-1.97)*** | 1.56 (1.35-1.81)*** | 1.61 (1.33-1.95)*** | Na | Na |
| **Num of BMI records** |  | 1.03 (1.03-1.03)*** | 1.02 (1.01-1.02)*** | 1.03 (1.03-1.03)*** | 1.02 (1.01-1.02)*** | 1.02 (1.02-1.03)*** | 1.01 (1.00-1.01)* | 1.02 (1.02-1.02)*** | 1.01 (1.00-1.01) | Na | Na |
| **BMI<27** |  | Reference | Reference | Reference | Reference | Reference | Reference | Reference | Reference | Na | Na |
| **Overweight** |  | Na | Na | 0.89 (0.70-1.13) | 1.01 (0.75-1.35) | 0.87 (0.68-1.11) | 0.99 (0.74-1.33) | 0.86 (0.67-1.09) | 0.99 (0.74-1.32) | Na | Na |
| **Obesity Class I** |  | Na | Na | 1.36 (1.08-1.71)** | 1.08 (0.81-1.44) | 1.24 (0.98-1.56) | 0.97 (0.73-1.30) | 1.24 (0.98-1.56) | 0.97 (0.72-1.29) | Na | Na |
| **Obesity Class II** |  | Na | Na | 1.90 (1.50-2.39)*** | 1.44 (1.07-1.93)* | 1.57 (1.24-1.98)*** | 1.19 (0.88-1.60) | 1.57 (1.24-1.99)*** | 1.19 (0.88-1.60) | Na | Na |
| **Obesity Class III** |  | Na | Na | 3.51 (2.78-4.43)*** | 2.05 (1.52-2.77)*** | 2.71 (2.14-3.44)*** | 1.62 (1.19-2.21)*** | 2.67 (2.11-3.39)*** | 1.60 (1.18-2.18)*** | Na | Na |
| **ASCVD** |  | Na | Na | Na | Na | 1.75 (1.59-1.94)*** | 2.37 (2.08-2.71)*** | 1.70 (1.54-1.88)*** | 2.34 (2.04-2.67)*** | Na | Na |
| **T2DM** |  | Na | Na | Na | Na | 1.26 (1.15-1.39)*** | 1.67 (1.47-1.91)*** | 1.27 (1.15-1.40)*** | 1.67 (1.47-1.91)*** | Na | Na |
| **HTN** |  | Na | Na | Na | Na | 1.79 (1.47-2.17)*** | 1.73 (1.34-2.25)*** | 1.62 (1.33-1.98)*** | 1.63 (1.26-2.12)*** | Na | Na |
| **HLP** |  | Na | Na | Na | Na | 0.91 (0.79-1.05) | 0.90 (0.74-1.10) | 0.92 (0.80-1.07) | 0.91 (0.75-1.12) | Na | Na |
| **CKD** |  | Na | Na | Na | Na | 1.56 (1.40-1.75)*** | 1.35 (1.15-1.58)*** | 1.52 (1.35-1.70)*** | 1.32 (1.12-1.55)*** | Na | Na |
| **NAFLD/NASH** |  | Na | Na | Na | Na | 1.18 (1.01-1.37)* | 0.94 (0.74-1.20) | 1.17 (1.01-1.37)* | 0.94 (0.74-1.19) | Na | Na |
| **OSA** |  | Na | Na | Na | Na | 1.31 (1.18-1.46)*** | 0.99 (0.85-1.16) | 1.32 (1.19-1.47)*** | 0.99 (0.85-1.16) | Na | Na |
| **SD of systolic BP** |  | Na | Na | Na | Na | Na | Na | 1.04 (1.04-1.05)*** | 1.03 (1.02-1.04)*** | Na | Na |
| **SD of HR** |  | Na | Na | Na | Na | Na | Na | 1.03 (1.01-1.04)*** | 1.03 (1.01-1.05)*** | Na | Na |
| **Mean BMI** |  | Na | Na | Na | Na | Na | Na | Na | Na | Na | Na |
| **Quartile of VIM of BMI** | Quartile2 | 1.05 (0.92-1.20) | 1.12 (0.94-1.34) | 1.09 (0.95-1.25) | 1.14 (0.95-1.36) | 1.08 (0.94-1.23) | 1.14 (0.95-1.36) | 1.05 (0.92-1.20) | 1.12 (0.93-1.33) | Na | Na |
|  | Quartile3 | 1.15 (1.00-1.31)* | 1.24 (1.04-1.49)* | 1.20 (1.05-1.37)** | 1.27 (1.06-1.52)*** | 1.17 (1.02-1.34)* | 1.25 (1.05-1.50)* | 1.12 (0.98-1.28) | 1.21 (1.01-1.45)* | Na | Na |
|  | Quartile4 | 1.41 (1.24-1.61)*** | 1.50 (1.25-1.79)*** | 1.48 (1.30-1.69)*** | 1.53 (1.28-1.83)*** | 1.40 (1.23-1.60)*** | 1.46 (1.22-1.76)*** | 1.28 (1.12-1.47)*** | 1.37 (1.14-1.65)*** | Na | Na |

Abbreviations: BMI, body mass index; HF, heart failure; HFpEF, heart failure with preserved ejection fraction; HFrEF, heart failure with rejected ejection fraction; SD, standard deviation; CV, coefficient of variation; VIM, variability independent of mean; ASV, average successive variability; ASCVD, atherosclerotic cardiovascular disease; T2DM, type 2 diabetes mellitus; HTN, hypertension; HLP, hyperlipidemia; CKD, chronic kidney disease; NAFLD/NASH, nonalcoholic fatty liver disease/nonalcoholic steatohepatitis; OSA, obstructive sleep apnea; SBP, systolic blood pressure; HR, heart rate.

Model 1 adjusted for age, gender, race, smoking status, and the number of BMI Records.

Model 2 adjusted for variables in model 1 plus baseline BMI classes.

Model 3: adjusted for variables in model 2 plus ASCVD, T2DM, HTN, HLP, CKD, NAFLD/NASH, and OSA.

Model 4: adjusted for variables in model 3 plus SD of SBP and SD of HR.

Model 5: adjusted for variables in model 4 plus mean BMI.

*p<0.05, **p<0.01, ***p<0.005

**Table S6. The Association between Clinical Characteristics with Incident Heart Failure by Quartiles of ASV of BMI**

| **Risk Factors** |  | **Models** | | | | | | | | | |
| --- | --- | --- | --- | --- | --- | --- | --- | --- | --- | --- | --- |
|  |  | **Model1** | | **Model2** | | **Model3** | | **Model4** | | **Model5** | |
| **HF Type** |  | HFpEF | HFrEF | HFpEF | HFrEF | HFpEF | HFrEF | HFpEF | HFrEF | HFpEF | HFrEF |
| **Age** |  | 1.08 (1.08-1.09)*** | 1.08 (1.08-1.09)*** | 1.09 (1.09-1.10)*** | 1.09 (1.08-1.09)*** | 1.07 (1.06-1.08)** | 1.07 (1.06-1.08)*** | 1.08 (1.07-1.08)*** | 1.07 (1.06-1.08)*** | 1.08 (1.07-1.08)*** | 1.07 (1.06-1.08)*** |
| **Gender** | Female | Reference | Reference | Reference | Reference | Reference | Reference | Reference | Reference | Reference | Reference |
|  | Male | 1.14 (1.04-1.25)** | 2.25 (1.98-2.56)*** | 1.26 (1.15-1.39)*** | 2.37 (2.09-2.70)*** | 1.02 (0.93-1.13) | 1.87 (1.64-2.13)*** | 1.04 (0.95-1.15) | 1.91 (1.67-2.18)*** | 1.06 (0.96-1.18) | 1.94 (1.70-2.22)*** |
| **Race** | White | Reference | Reference | Reference | Reference | Reference | Reference | Reference | Reference | Reference | Reference |
|  | Black | 1.15 (1.00-1.32) | 1.36 (1.12-1.65)*** | 1.09 (0.95-1.26) | 1.32 (1.09-1.61)** | 1.00 (0.86-1.15) | 1.15 (0.95-1.41) | 0.95 (0.82-1.10) | 1.12 (0.92-1.36) | 0.94 (0.81-1.08) | 1.11 (0.91-1.35) |
|  | Other | 1.16 (0.76-1.77) | 1.73 (1.43-2.08) | 1.23 (0.81-1.88) | 1.48 (0.87-2.52) | 1.27 (0.83-1.93) | 1.54 (0.91-2.61) | 1.26 (0.83-1.92) | 1.54 (0.91-2.62) | 1.28 (0.84-1.95) | 1.56 (0.92-2.64) |
| **Smoking status** |  | 1.54 (1.33-1.78)*** | 1.73 (1.43-2.08)*** | 1.67 (1.44-1.93)*** | 1.79 (1.48-2.16)*** | 1.60 (1.39-1.85)*** | 1.65 (1.37-2.00)*** | 1.57 (1.35-1.81)*** | 1.63 (1.35-1.97)*** | 1.57 (1.36-1.81)*** | 1.63 (1.35-1.97)*** |
| **Num of BMI records** |  | 1.03 (1.03-1.04)*** | 1.02 (1.02-1.03)*** | 1.03 (1.03-1.04)*** | 1.02 (1.02-1.03)*** | 1.02 (1.02-1.03)*** | 1.01 (1.00-1.02)*** | 1.02 (1.02-1.03)*** | 1.01 (1.00-1.01)* | 1.02 (1.02-1.03)*** | 1.01 (1.00-1.01)* |
| **BMI<27** |  | Reference | Reference | Reference | Reference | Reference | Reference | Reference | Reference | Reference | Reference |
| **Overweight** |  | Na | Na | 0.84 (0.66-1.07) | 0.95 (0.71-1.27) | 0.83 (0.66-1.06) | 0.94 (0.71-1.26) | 0.83 (0.65-1.05) | 0.95 (0.71-1.26) | 0.80 (0.62-1.01) | 0.91 (0.68-1.21) |
| **Obesity Class I** |  | Na | Na | 1.24 (0.99-1.56) | 0.98 (0.74-1.31) | 1.14 (0.91-1.44) | 0.90 (0.67-1.20) | 1.16 (0.92-1.46) | 0.91 (0.68-1.21) | 0.98 (0.77-1.24) | 0.76 (0.56-1.04) |
| **Obesity Class II** |  | Na | Na | 1.63 (1.29-2.05)*** | 1.25 (0.93-1.67) | 1.37 (1.08-1.73)** | 1.05 (0.78-1.41) | 1.42 (1.12-1.79)*** | 1.08 (0.80-1.45) | 1.01 (0.77-1.32) | 0.76 (0.53-1.10) |
| **Obesity Class III** |  | Na | Na | 2.84 (2.25-3.60)*** | 1.71 (1.26-2.32)*** | 2.24 (1.77-2.85)*** | 1.38 (1.01-1.88)* | 2.32 (1.83-2.95)*** | 1.41 (1.03-1.92)* | 1.71 (1.55-1.89) | 0.77 (0.47-1.25) |
| **ASCVD** |  | Na | Na | Na | Na | 1.76 (1.59-1.94)*** | 2.38 (2.08-2.72)*** | 1.70 (1.55-1.88)*** | 2.34 (2.05-2.68)*** | 1.71 (1.55-1.89)*** | 2.35 (2.06-2.69)*** |
| **T2DM** |  | Na | Na | Na | Na | 1.27 (1.15-1.39)*** | 1.68 (1.47-1.92)*** | 1.27 (1.15-1.40)*** | 1.68 (1.47-1.91)*** | 1.24 (1.12-1.36)*** | 1.64 (1.44-1.87)*** |
| **HTN** |  | Na | Na | Na | Na | 1.78 (1.47-2.17)*** | 1.74 (1.34-2.25)*** | 1.62 (1.33-1.97)*** | 1.64 (1.26-2.13)*** | 1.59 (1.31-1.94)*** | 1.61 (1.24-2.10)*** |
| **HLP** |  | Na | Na | Na | Na | 0.91 (0.79-1.05) | 0.90 (0.73-1.10) | 0.92 (0.80-1.07) | 0.91 (0.75-1.12) | 0.93 (0.80-1.07) | 0.91 (0.75-1.12) |
| **CKD** |  | Na | Na | Na | Na | 1.57 (1.40-1.76)*** | 1.36 (1.16-1.60)*** | 1.52 (1.36-1.70)*** | 1.33 (1.13-1.56)*** | 1.52 (1.36-1.70)*** | 1.33 (1.13-1.56)*** |
| **NAFLD/NASH** |  | Na | Na | Na | Na | 1.17 (1.01-1.37)* | 0.94 (0.74-1.19) | 1.17 (1.00-1.37)* | 0.94 (0.74-1.19) | 1.17 (1.00-1.36)* | 0.93 (0.74-1.19) |
| **OSA** |  | Na | Na | Na | Na | 1.31 (1.18-1.45)*** | 0.99 (0.85-1.15) | 1.32 (1.18-1.47)*** | 0.99 (0.85-1.15) | 1.28 (1.15-1.43)*** | 0.97 (0.83-1.13) |
| **SD of systolic BP** |  | Na | Na | Na | Na | Na | Na | 1.04 (1.04-1.05)*** | 1.03 (1.02-1.04)*** | 1.05 (1.04-1.05)*** | 1.03 (1.02-1.04)*** |
| **SD of HR** |  | Na | Na | Na | Na | Na | Na | 1.02 (1.01-1.04)*** | 1.03 (1.01-1.05)*** | 1.02 (1.01-1.04)*** | 1.03 (1.01-1.05)*** |
| **Mean BMI** |  | Na | Na | Na | Na | Na | Na | Na | Na | 1.04 (1.02-1.06)*** | 1.04 (1.02-1.06)*** |
| **Quartile of ASV of BMI** | Quartile2 | 1.33 (1.16-1.52) | 1.09 (0.92-1.30) | 1.16 (1.01-1.33)* | 1.04 (0.87-1.24) | 1.13 (0.99-1.30) | 1.02 (0.86-1.22) | 1.11 (0.97-1.28) | 1.01 (0.85-1.20) | 1.11 (0.97-1.27) | 1.00 (0.84-1.20) |
|  | Quartile3 | 1.59 (1.39-1.82)*** | 1.37 (1.15-1.64)*** | 1.26 (1.09-1.44)*** | 1.24 (1.04-1.48)* | 1.23 (1.07-1.41)*** | 1.24 (1.03-1.48)* | 1.17 (1.02-1.35)* | 1.19 (1.00-1.43) | 1.16 (1.00-1.33)* | 1.18 (0.98-1.41) |
|  | Quartile4 | 2.26 (1.98-2.57)*** | 1.63 (1.37-1.93)*** | 1.56 (1.36-1.79)*** | 1.36 (1.13-1.63)*** | 1.49 (1.29-1.71)*** | 1.32 (1.09-1.58)*** | 1.35 (1.18-1.56)*** | 1.23 (1.02-1.48)* | 1.29 (1.12-1.49)*** | 1.17 (0.97-1.42) |

Abbreviations: BMI, body mass index; HF, heart failure; HFpEF, heart failure with preserved ejection fraction; HFrEF, heart failure with rejected ejection fraction; SD, standard deviation; CV, coefficient of variation; VIM, variability independent of mean; ASV, average successive variability; ASCVD, atherosclerotic cardiovascular disease; T2DM, type 2 diabetes mellitus; HTN, hypertension; HLP, hyperlipidemia; CKD, chronic kidney disease; NAFLD/NASH, nonalcoholic fatty liver disease/nonalcoholic steatohepatitis; OSA, obstructive sleep apnea; SBP, systolic blood pressure; HR, heart rate.

Model 1 adjusted for age, gender, race, smoking status, and the number of BMI Records.

Model 2 adjusted for variables in model 1 plus baseline BMI classes.

Model 3: adjusted for variables in model 2 plus ASCVD, T2DM, HTN, HLP, CKD, NAFLD/NASH, and OSA.

Model 4: adjusted for variables in model 3 plus SD of SBP and SD of HR.

Model 5: adjusted for variables in model 4 plus mean BMI.

*p<0.05, **p<0.01, ***p<0.005

**Table S7. The Association between BMI Variability with Heart Failure by Quartiles of BMI Variability with Maximum of 7.5 Years of Follow-up**

| **Measurement of BMI Variability** | **Quartiles of BMI variability** | | | | | | | | **CV of BMI** | |
| --- | --- | --- | --- | --- | --- | --- | --- | --- | --- | --- |
| **HF Subtype** | **HFpEF** | | | | **HFrEF** | | | | **HFpEF** | **HFrEF** |
|  | Q1 | Q2 | Q3 | Q4 | Q1 | Q2 | Q3 | Q4 | 1-SD increment | 1-SD increment |
| **SD** | Reference | 1.13 (0.97-1.31) | 1.17 (1.01-1.35)* | 1.34 (1.14-1.56)*** | Reference | 1.07 (0.89-1.29) | 1.26 (1.04-1.52)* | 1.42(1.16-1.75)*** | 1.10 (1.04-1.17)*** | 1.10 (1.01-1.19)* |
| **CV** | Reference | 1.02 (1.89-1.18) | 1.05 (0.92-1.21) | 1.25 (1.08-1.43)*** | Reference | 1.16 (0.97-1.39) | 1.26 (1.05-1.51)* | 1.40 (1.15-1.69)*** | 1.04 (1.02-1.06)*** | 1.04 (1.01-1.07)* |
| **VIM** | Reference | 1.06 (0.92-1.21) | 1.10 (0.96-1.26) | 1.29 (1.13-1.48)*** | Reference | 1.06 (0.88-1.27) | 1.19 (0.99-1.43） | 1.36 (1.13-1.63)*** | 1.17 (1.06-1.28)*** | 1.17 (1.03-1.33)* |
| **ASV** | Reference | 1.11 (0.96-1.28) | 1.15 (1.00-1.33) | 1.30 (1.12-1.50)*** | Reference | 0.98 (0.82-1.17) | 1.13 (0.94-1.36) | 1.16 (0.96-1.40) | 1.13 (1.04-1.30)*** | 1.09 (0.96-.1.23) |

Abbreviations: BMI, body mass index; HF, heart failure; HFpEF, heart failure with preserved ejection fraction; HFrEF, heart failure with rejected ejection fraction; SD, standard deviation; CV, coefficient of variation; VIM, variability independent of mean, ASV, average successive variability.

Adjusted for all covariates in model 5 (in model 4 for VIM).

*p<0.05, **p<0.01, ***p<0.005.
